# Discordance between source and point-of-use drinking-water *Escherichia coli* contamination in Bangladesh: a cross-sectional analysis and implications for safely managed water monitoring

**DOI:** 10.64898/2026.09.29.26364109

**Authors:** S. M. Ashraf Ul Hoque, Md Khairul Islam, Mohammad Khalid Hasan, Mohiuddin Sharif, Bilkis Banu, Zenat Zebin Hossain, Kamran ul Baset

## Abstract

**Background:** Safely managed drinking-water monitoring under Sustainable Development Goal target 6.1 uses source-water quality, yet *Escherichia coli* is common in household drinking water in Bangladesh despite near-universal improved sources. We compared source and household point-of-use (POU) *E. coli* detection within households and estimated how the safely managed estimate changes if POU replaces source *E. coli*.

**Methods:** Cross-sectional secondary analysis of the Bangladesh MICS 2025 water-quality subsample (February–June 2025; detection ≥1 colony-forming unit/100 mL). We classified 5,944 households with readable paired results into four joint patterns. As secondary aims, survey-weighted modified Poisson models examined POU detection among negative-source households and two-week diarrhoea in children under five.

**Results:** *E. coli* was detected in 47.4% of source and 84.8% of POU samples; only 12.7% of households were negative at both. Among negative-source households, 75.8% had *E. coli* in household water, versus 94.7% with a detected source, persisting (33.3%) even direct from the source. The JMP-aligned safely managed estimate was 42.5% (95% CI 40.8–44.1); substituting POU for source *E. coli* gave 12.6% (11.4–13.8), not an SDG indicator estimate. The diarrhoea prevalence ratio for POU detection (1.04, 0.59–1.86) was too imprecise to interpret. In exploratory spatial analysis, POU detection clustered geographically (Moran’s I 0.16 cluster level, 0.31 district level, both p<0.001) and was independently associated with lower elevation (odds ratio 0.62, 0.56–0.70) and longer travel time to healthcare (1.65, 1.29–2.12); five districts had complete detection among 84–94 households tested.

**Conclusions:** Source and POU classifications differed substantially; the survey cannot show where or when contamination occurred. Contamination was spatially clustered, arguing against pure noise and pointing to districts for follow-up. The findings support evaluating standardised POU sampling as a complement to source-based monitoring, with targeted follow-up and cross-season, cross-country replication.

## Introduction

Diarrhoeal disease continues to cause illness and death in children under five in South Asia and sub-Saharan Africa [1,2], and inadequate water, sanitation and hygiene were estimated to account for 60% of diarrhoeal deaths in 2016 [3]. Under Sustainable Development Goal (SDG) target 6.1, safely managed drinking water requires an improved source on premises, available when needed and free of faecal contamination at the source. In 2024, 2.1 billion people still lacked such services [4]; in a geospatial analysis of 135 low- and middle-income countries, faecal contamination was the main limiting factor [5]. The World Health Organization (WHO) guideline is that *Escherichia coli* (*E. coli*) should not be detectable in any 100 mL sample of drinking water [6]. The WHO/UNICEF Joint Monitoring Programme (JMP) core survey questions include an *E. coli* test of source water (question W6), with household-water testing and observation of storage containers in the expanded question list [7].

The Bangladesh Multiple Indicator Cluster Survey (MICS) 2025 found that 99.0% of the population used an improved source and 98.6% at least basic drinking-water services. *E. coli* was detected in source water used by 47.1% of household members and in household drinking water for 85.0%; 42.6% met the MICS definition of safely managed drinking water [8].

Prior studies suggest that water quality often deteriorates between source and home. In Sierra Leone all improved-source samples met the WHO guideline while 92.9% of stored household samples contained thermotolerant coliforms [9]; in Rohingya camps in Bangladesh 74% of stored samples were faecally contaminated compared with 28% of tubewell samples [10]. A systematic review of 57 studies found that bacteriological quality declines after collection in many settings [11]. A meta-analysis of 45 studies found mean non-compliance of 46% at source and 75% in stored water [12], and in rural India 69% of stored samples contained *E. coli* [13]. More than a quarter of samples from improved sources were faecally contaminated in 38% of 191 studies [14].

Across 27 MICS surveys, *E. coli* was found at the point of collection in 16–90% of households and at the point of use (POU) in 19–99%, with more POU contamination in rural and wet-season samples [15]. That multi-country analysis compared point-of-collection and POU prevalence across surveys but did not classify individual households by their joint result or recalculate the safely managed indicator under a POU-substituted definition. In Bangladesh, two analyses of MICS 2019 examined POU *E. coli* determinants [16] and its association with under-five diarrhoea [17]. Those Bangladeshi analyses used household drinking water alone. To our knowledge, no national analysis has classified households by the combination of source and POU results or examined what that implies for the safely managed estimate.

MICS 2025 tested source and household drinking water in the same households, which allows a direct within-household comparison. Therefore, our aim was to classify households in Bangladesh by joint source and POU *E. coli* status, and to assess how this changes the safely managed drinking-water estimate. As secondary and exploratory aims, we examined factors associated with POU detection in negative-source households, joint patterns and two-week diarrhoea in children under five, and geographic clustering of POU detection.

## Materials and methods

### Data and study design

We analysed Bangladesh MICS 2025 microdata (Bangladesh Bureau of Statistics with UNICEF), a two-stage stratified cluster survey with 3,149 primary sampling units (PSUs) from the 2022 census frame and 20 households per PSU (62,980 sampled; 61,207 interviewed). Water-quality testing was done in two randomly chosen households per PSU (6,298 sampled). Blank controls were run for one in five clusters. The microdata were obtained under the MICS data-use terms and are not redistributed. The analysis followed the STROBE guidance [18]; the checklist is in S1 STROBE Checklist.

### Ethics statement

According to the survey report, the MICS 2025 survey plan, questionnaires and protection protocol were approved by the icddr,b Ethical Review Committee in November 2024 (reference PR-24113), and verbal informed consent was obtained from respondents [8]. For water testing, the household respondent was asked for verbal permission, recorded on the survey tablet, and was told that individual results would not be returned to households. This secondary analysis used only anonymised MICS microdata obtained under the MICS data-use terms; the authors had no contact with participants or access to identifying information. Because ethical review and informed consent were obtained by the survey implementers, the authors did not seek separate ethical approval.

### Analysis hierarchy

Analyses were organised into three levels. Primary: source and POU detection; the four-category joint distribution and the asymmetry of the discordant patterns; and the JMP-aligned and POU-substituted safely managed estimates. Secondary: factors associated with POU detection among negative-source households, and joint patterns and diarrhoea. Exploratory: spatial clustering and geospatial covariates, divisional patterns, and sensitivity analyses. The sensitivity analyses covered the ≥11 CFU/100 mL threshold, risk groups, source-by-container interaction, survey month and other checks (in S1 File where not shown in the main text).

### Water testing

#### Collection

Interviewers asked for a glass of the water that household members usually drink and recorded whether it was poured directly from the source or from a covered or uncovered container. Where possible they also sampled the household’s main source. Both samples were collected in Whirl-Pak bags and tested within 30 minutes (Bangladesh MICS 2025 water-quality testing form [8]).

#### Laboratory method

The MICS method filters 100 mL through a 0.45 µm membrane placed on a Compact Dry EC plate [15]; blue colonies were counted after 24–36 hours of incubation [8]. Counts are per 100 mL; Counts above the upper limit of the test were truncated at the maximum readable value, and unreadable or lost results were set to missing. No observations were removed as outliers. Detection was ≥1 colony-forming unit (CFU)/100 mL.

#### Quality control

Blank controls used mineral water in 10% of sampled households (one household in every fifth cluster); 7 of 605 (1.2%) had *E. coli* detected. Blanks test the filtration and laboratory steps, not the glass, hands or pouring. Measurers received 15 days of water-quality training including a pilot, and 43 of 45 trained were selected on performance [8].

#### Not recorded

The Bangladesh form gives no instruction to flush the source before sampling. The generic MICS manual states that point-of-collection samples should be flushed for 30 seconds whenever feasible (tube wells pumped, taps opened) [19]; the report does not say which manual was used or document adherence, so we cannot confirm flushing. The data also do not record how long the water in the glass had been stored, whether the glass was clean, or how much time elapsed between the two collections. The Water Quality Testing Form fixes the collection sequence: household glass water is collected first, and the source sample second [8]. Because of this fixed order, any post-collection handling of the source sample cannot explain contamination detected in the household glass, although contamination of the glass before the visit remains possible. The glass water need not come from the sampled source: the recorded source of the glass differs from the main source code in 17% of households.

#### Timing

Fieldwork ran from 24 February to 30 June 2025 according to the report; two water tests are dated July. Testing was concentrated in March to June (95%), with 649 of the 6,106 tested households in June or July.

### Joint status

Among households with readable results at both locations we defined four exclusive patterns, following the point-of-collection/point-of-use cross-tabulation used in the multi-country MICS analysis of Bain et al. [15]: not detected at either location (ND/ND); source ND, POU detected (ND/Detected); source detected, POU ND (Detected/ND); detected at both. Both samples come from one visit, so the patterns describe a point-in-time difference between two samples. We report the survey-weighted difference between the two discordant shares, with a design-based confidence interval, and their ratio as descriptive measures. Source and POU samples come from different locations and procedures, so this asymmetry is not a test of sampling error.

### JMP-aligned and POU-substituted estimates of safely managed drinking water

We reproduced the JMP-aligned MICS definition [7,8]: improved source (piped water, tube wells, protected wells and springs, rainwater, delivered or packaged water), on premises (in the dwelling or yard), sufficient water when needed in the past month, and no *E. coli* in the source sample. JMP defines on-premises access by location rather than a fixed distance threshold, and MICS does not record the physical distance between the point of use and the source. The JMP definition also requires freedom from priority chemical contamination; the MICS indicator applies *E. coli* only, and so do we [7,8]. We formed a POU-substituted estimate by replacing the source criterion with no *E. coli* in the POU sample, and a version requiring both. The POU-substituted estimate is a monitoring counterfactual; the SDG indicator is defined by source-water quality and is unchanged. These estimates refer to the household population (weight multiplied by household size), among households with readable paired results; prevalence and joint-pattern estimates are household-weighted. We validated the JMP-aligned estimate against the official report (Results); on-premises water use was 85.5% in the paired sample (85.6% among all 6,010 source-tested households) against the official 85.9%.

### Exposure and covariate definitions

Storage container, household water treatment, handwashing place, collection time, source type and sanitation facility type follow the JMP core and expanded household-survey question definitions [7]: storage container was the observed vessel from which the glass was taken (direct from source, covered container or uncovered container; covered container was the reference, so the direct-from-source group was analysed as its own category); household water treatment was reported at the test visit and counted as effective if the household boiled, chlorinated, filtered or solar-disinfected water, and not if it only strained or settled the water; handwashing place was an observed place with water and soap or detergent; collection time was a round trip of ≥30 minutes versus on-premises or shorter; sanitation was classified as an improved or unimproved facility type. Wealth quintile used the standard DHS asset-based wealth index [20]. Other covariates were residence, division, head’s education (four levels) and survey month (grouped); for children, age group, sex and mother’s education.

Definitions were set after inspecting how variables were coded and before outcome models were run.

### Spatial data and geospatial covariates (exploratory)

We linked each water-quality cluster to its GPS coordinates using the MICS 2025 GPS dataset. Following standard DHS practice [21,22], UNICEF displaces MICS cluster coordinates for respondent confidentiality (typically up to 2 km for urban clusters and 5 km for rural clusters, with 1% of rural clusters displaced up to 10 km, constrained not to cross an administrative boundary); in this survey the accompanying documentation states that clusters were displaced within the third administrative level (upazila), so coordinates describe the local area, not exact household locations. Point-extracted covariates (for example elevation and travel time) therefore carry additional positional error beyond the ecological-aggregation error already noted for the district-level model (Strengths and limitations). The companion Geospatial Covariates (GeoCov) dataset provides cluster- and district-level values for environmental, climate, population and accessibility variables derived from external raster and administrative sources, including elevation, CHIRPS-derived rainfall (2020 layer, the closest available year to the 2025 survey in the standard GeoCov release), Gridded Population of the World v4 (GPWv4) population density (2020), a Malaria Atlas Project motorised travel-time-to-healthcare surface (2019) and a VIIRS nighttime-lights composite; exact source datasets, spatial resolutions and vintages for each covariate are listed in S1 File.

### Statistical analysis

#### Design and general approach

All analyses accounted for the MICS survey design: PSU and stratum variables, Taylor-series variance (singleton strata centred), and the water-quality weight for the sample analysed (one weight for paired and child analyses, a separate weight for POU-only estimates). Confidence intervals and p-values were two-sided (α = 0.05); CIs for proportions are logit-based. No sample-size calculation was done; sample size followed the survey design. No multiplicity correction was applied, because the secondary and exploratory analyses (factors associated with POU detection, diarrhoea and spatial clustering) are hypothesis-generating and no conclusion rests on a single test. Subgroup and sensitivity comparisons within these analyses (for example by division, source type or container) were likewise treated as descriptive, not confirmatory. The plan was not preregistered; the full-sample model, the weighting analysis, the direct-from-source benchmark, the spatial analysis and several sensitivity analyses were added after the first results were seen. Missing data were handled by complete-case analysis, which kept at least 96% of eligible households and children, so we did not impute (S2 File).

The remaining procedures are grouped by study objective. Each paragraph states only how that objective’s outcome was analysed; full model specifications, sensitivity analyses and diagnostics are in S1 File and S2 File.

#### Detection, joint patterns and safely managed estimates

We estimated source and POU detection, the four joint patterns, and the JMP-aligned and POU-substituted safely managed indicators as survey-weighted proportions with design-based confidence intervals. The discordant-pattern asymmetry was tested as a weighted difference in shares. Four sensitivity analyses in S1 File each address one threat. Excluding clusters with a positive blank control addresses laboratory contamination. Inverse-probability-of-observation weighting addresses selection bias from unreadable pairs. Restricting to households whose glass source matched the main source addresses source misclassification. Excluding delivered or packaged water addresses a different supply chain.

#### Factors associated with POU detection

Among households with a negative source, modified Poisson regression (log link, robust variance) estimated prevalence ratios for storage, treatment, handwashing, collection time and wealth. We used modified Poisson regression because POU detection exceeded 75% and odds ratios would overstate prevalence ratios. Covariates were chosen on substantive grounds as likely common causes of household water handling and POU contamination, as shown in the directed acyclic graph (S1 Fig). Two adjustment sets were used: minimal (wealth, residence, division, head’s education) and mutual (adding all handling factors, source type and sanitation). Estimates were similar under the minimal and mutual adjustment sets (Table 4). A parallel model in all paired households added source status and source-by-container interaction terms, reported as prevalence ratios with 95% confidence intervals. These associations are descriptive, not causal, because treatment and storage may respond to perceived water quality. S1 File reports adjusted prevalence differences and further sensitivity analyses. Restricting to stored water addresses the different container reference group. Restricting to on-premises sources addresses contamination during collection. Excluding households that treated water addresses reverse causation. Adding survey month addresses seasonal confounding. A ≥11 CFU/100 mL threshold addresses misclassification near the detection limit.

#### Childhood diarrhoea

Modified Poisson models compared two-week diarrhoea by POU detection and, separately, by joint pattern (reference ND/ND), adjusting for child age, sex, mother’s education, wealth, residence and sanitation, with PSU-clustered variance. Children carried the water-quality weight of their household, which reflects the selection of two households per PSU for testing. Minimum detectable PRs assumed 80% power, α = 0.05 and a design effect of 1.30, estimated from the weighted diarrhoea prevalence. Sensitivity analyses added survey month, handwashing place and treatment, to address season and other transmission routes. S1 File reports further checks using the under-five child weight, minimal adjustment, on-premises sources, treatment exclusion and a ≥11 CFU/100 mL threshold.

#### Spatial clustering

Cluster and district GPS coordinates [23] were linked to POU detection. Global Moran’s I [24,25] tested spatial autocorrelation at both levels. A household-level mixed-effects logistic model incorporating the household survey weight via multilevel pseudo maximum likelihood [26] and a district-level ecological model, weighted by the number of households tested, tested association with geospatial covariates. Because POU detection exceeded 80%, odds ratios from this model overstate the corresponding prevalence ratios; we therefore interpret them for direction, not effect size [27]. Inverse-variance weights are undefined for districts with complete detection, so inverse-variance weighting and exclusion of those districts were sensitivity analyses (S1 File). The household model used two levels (household in PSU): MICS’s primary sampling unit is the PSU, not the district, and no separate district-level survey weight exists, so district was not added as a further weighted level; independent evidence on district-level clustering comes from the ecological model and the polygon-adjacency Moran’s I. Full specifications are in S1 File; the linkage, displacement and covariate-provenance details are in Materials and methods, “Spatial data and geospatial covariates (exploratory)”.

#### Software

Survey-weighted models were fitted in Stata 17 (StataCorp, College Station, Texas, USA). Spatial analyses, figures and a cross-check of the Stata estimates used R 4.6.1 with the packages survey, spdep, WeMix and sf.

## Results

### Sample

Of 6,106 households with a completed POU test, 6,048 had a readable POU count; source samples were collected in 6,010 and readable in 5,999; both were readable in 5,944 (the paired sample; S2 Fig). The 162 households lost from the paired sample were less often on premises (61.8% versus 85.2%) and more often used other source types (23.0% versus 2.2%) than included households (S2 File), so the estimates describe households with readable paired results. Weighting for the probability of readable paired results changed the estimates little (source detection 47.5%, POU detection 84.8%, JMP-aligned estimate 42.2%, POU-substituted estimate 12.5%; S1 File).

Of 2,442 children under five in tested households (more than one child per household in some cases, so household and child counts are not identical), 2,377 lived in paired households. Of these, 126 were excluded because their questionnaire was not completed and no age was recorded, leaving 2,251 aged 0–59 months; 2,249 of them had valid diarrhoea and covariate data (1,727 PSUs; 1,596 degrees of freedom). Table 1 describes the paired sample and S2 File gives the counts excluded at each step.

**Table 1.** Characteristics of the paired analytic sample (weighted %).

| Characteristic | Paired sample (n = 5,944) |
| --- | --- |
| **Residence** |  |
| Urban | 30.2 |
| Rural | 69.8 |
| **Wealth quintile** |  |
| Poorest | 21.0 |
| Second | 19.8 |
| Middle | 19.5 |
| Fourth | 19.7 |
| Richest | 20.1 |
| **Division** |  |
| Barishal | 5.3 |
| Chattogram | 18.2 |
| Dhaka | 26.9 |
| Khulna | 11.6 |
| Mymensingh | 7.4 |
| Rajshahi | 13.8 |
| Rangpur | 11.3 |
| Sylhet | 5.4 |
| **Household head's education** |  |
| None/pre-primary | 29.9 |
| Primary | 25.6 |
| Secondary | 28.6 |
| Higher secondary or above | 15.9 |
| **Main source type** |  |
| Piped | 25.6 |
| Deep tube well | 20.5 |
| Shallow tube well | 51.5 |
| Other | 2.3 |
| **Container at POU test** |  |
| Direct from source | 14.5 |
| Covered container | 67.8 |
| Uncovered container | 17.7 |
| **Household water treatment** |  |
| Effective treatment reported | 7.2 |
| No effective treatment | 92.8 |
| **Handwashing place** |  |
| Handwashing place with water and soap | 68.6 |
| Not observed | 31.4 |
| **Collection round trip** |  |
| On premises or <30 min | 98.8 |
| ≥30 min | 1.2 |
| **Sanitation** |  |
| Improved sanitation | 89.7 |
| Unimproved sanitation | 10.3 |

*Percentages within each characteristic are weighted; container rows are based on the 5,806*
*paired households with an observed container.*

### Source and POU detection

*E. coli* was detected in 47.4% (95% CI 45.9–49.0) of source samples and 84.8% (83.6–85.9) of POU samples in the paired sample (Table 2). On the full tested samples the estimates were 47.6% (46.0–49.1; n = 5,999) and 84.5% (83.3–85.6; n = 6,048). POU detection was lower in urban (78.2%) than rural (87.7%) households and in the richest quintile (74.5%) than in the other four (84.7–89.8%). Source detection ranged from 29.7% in Mymensingh and 30.6% in Rangpur to 63.5% in Khulna.

**Table 2.** *E. coli* detection (≥1 CFU/100 mL) in source and household POU water, paired sample, MICS Bangladesh 2025.

| Subgroup | Source, % (95% CI) | POU, % (95% CI) | Unweighted n |
| --- | --- | --- | --- |
| **National** | **47.4 (45.9–49.0)** | **84.8 (83.6–85.9)** | **5,944** |
| Urban | 46.3 (43.3–49.4) | 78.2 (75.6–80.6) | 1,609 |
| Rural | 47.9 (46.1–49.7) | 87.7 (86.4–88.8) | 4,335 |
| Poorest | 47.0 (44.0–49.9) | 88.0 (86.0–89.8) | 1,455 |
| Second | 47.3 (44.2–50.5) | 89.8 (87.7–91.6) | 1,269 |
| Middle | 51.1 (47.7–54.5) | 87.0 (84.4–89.3) | 1,178 |
| Fourth | 48.0 (44.4–51.6) | 84.7 (81.8–87.2) | 1,074 |
| Richest | 43.9 (40.3–47.6) | 74.5 (71.2–77.6) | 968 |
| Barishal | 38.1 (33.9–42.6) | 91.0 (88.0–93.4) | 524 |
| Chattogram | 50.1 (46.0–54.3) | 88.3 (85.8–90.4) | 1,047 |
| Dhaka | 48.5 (45.3–51.8) | 80.6 (77.8–83.2) | 1,354 |
| Khulna | 63.5 (59.7–67.2) | 93.7 (91.5–95.4) | 860 |
| Mymensingh | 29.7 (24.2–35.8) | 66.2 (59.3–72.4) | 345 |
| Rajshahi | 51.0 (46.9–55.1) | 88.3 (85.2–90.8) | 731 |
| Rangpur | 30.6 (26.9–34.6) | 83.6 (80.6–86.3) | 728 |
| Sylhet | 57.5 (52.0–62.7) | 87.9 (83.5–91.2) | 355 |

*Survey-weighted; n is the unweighted number of paired households.*

### Joint patterns and discordance

Only 12.7% of households had no detectable *E. coli* at either location (Table 3; Fig 1), and this was more common in urban households (17.8%) and the richest quintile (20.3%). Among the 3,063 households with a negative source, 75.8% (73.9–77.7) had *E. coli* in POU water, compared with 94.7% (93.6–95.7) of the 2,881 households with a detected source. The two discordant patterns were very unequal: 2,372 households were ND/Detected and 128 Detected/ND (weighted 39.9% versus 2.5%; difference 37.4 percentage points, 95% CI 35.7–39.0; ratio 16.0, 12.6–19.5).

**Fig 1.**
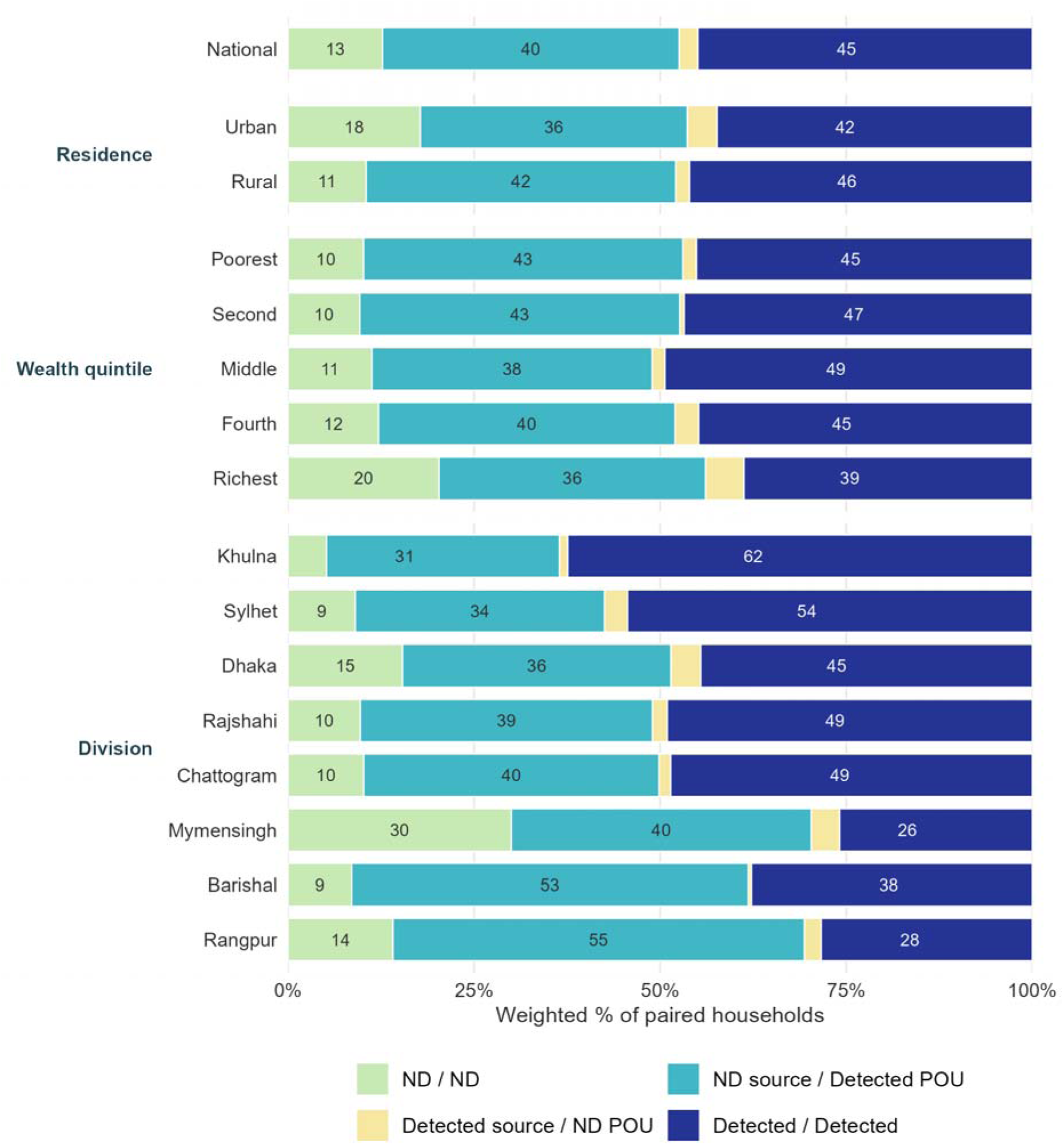
Joint source and POU *E. coli* status. Weighted percentage of paired households in the four joint patterns, nationally and by residence, wealth quintile and division (ND, not detected). The two samples differ in location, timing and handling, so the patterns do not identify when or where contamination occurred.

**Table 3.** Joint source–POU *E. coli* status, paired sample (n = 5,944).

| Pattern (source / POU) | Unweighted n | Weighted % (95% CI) |
| --- | --- | --- |
| ND / ND | 691 | 12.7 (11.7–13.8) |
| Source ND / POU detected | 2,372 | 39.9 (38.4–41.4) |
| Source detected / POU ND | 128 | 2.5 (2.0–3.1) |
| Detected / detected | 2,753 | 44.9 (43.4–46.4) |

Water storage does not fully explain the discordance. ND/Detected was 33.3% (29.5–37.4) when the glass was poured directly from the source, 39.0% for covered containers and 48.3% for uncovered containers. Discordance therefore persisted with almost no storage time. By source type, ND/Detected was 36.4% for piped supply, 42.0% for deep tube wells, 41.2% for shallow tube wells and 31.0% for other sources, and POU detection among source-negative households was 66.1%, 82.8%, 78.3% and 74.5% (n = 53 for other sources).

Excluding the 13 paired households in the seven clusters with a positive blank control, restricting to the 4,963 households in which the glass source code matched the main source code, and excluding households using delivered or packaged water left the estimates essentially unchanged (ND/Detected 39.8–39.9%; JMP-aligned 42.1–42.5%; POU-substituted 12.5–12.6%; S1 File). Source detection was 41.3% in February–March, 50.1% in April, 51.2% in May and 50.7% in June–July; POU detection was 80.7%, 85.4%, 86.8% and 91.8%. Fig 2 shows the joint distribution by contamination class.

**Fig 2.**
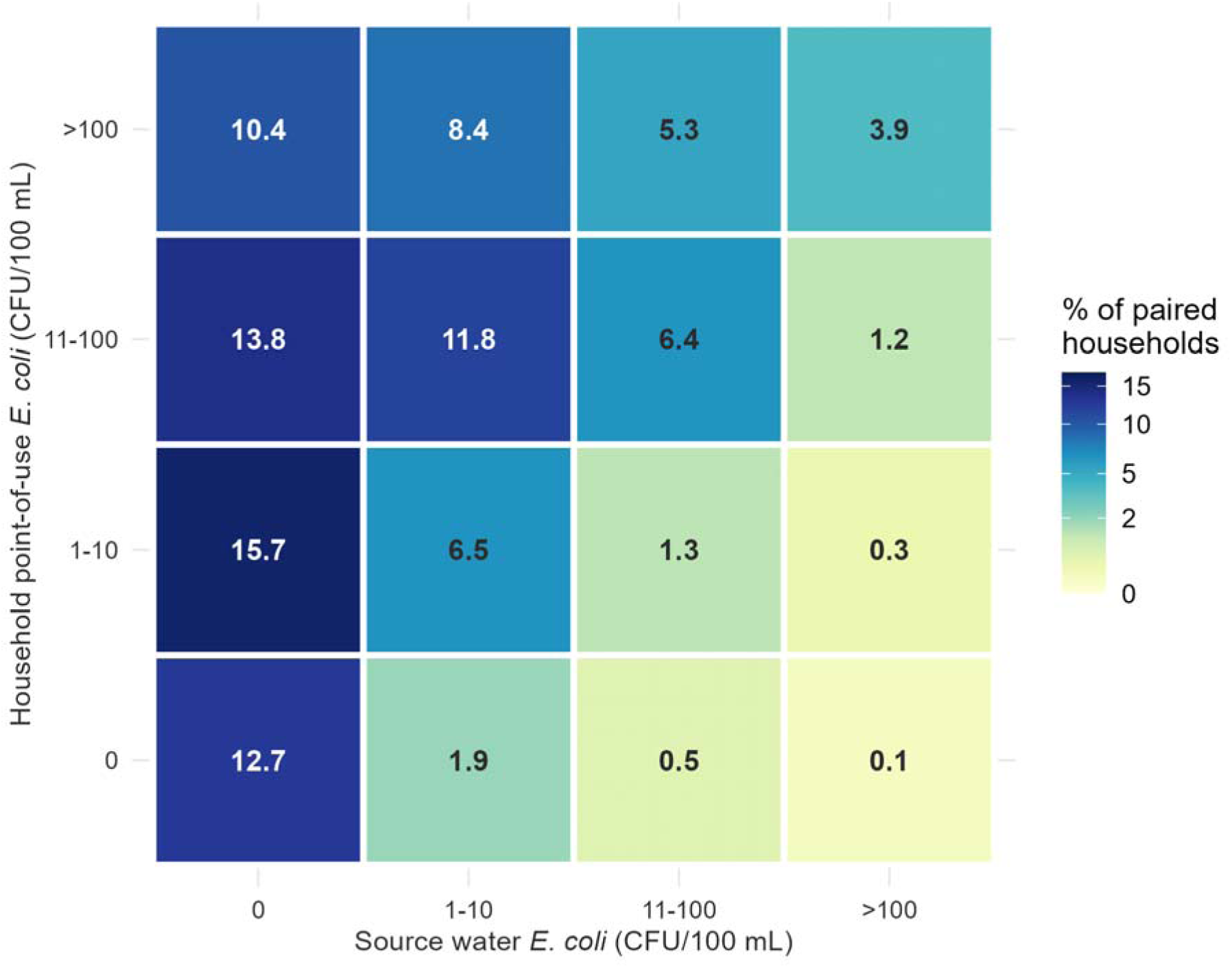
Concordance matrix of source and POU *E. coli* counts. Weighted percentage of paired households by count class (0, 1–10, 11–100, >100 CFU/100 mL) at the source and at the point of use.

### JMP-aligned and POU-substituted estimates of safely managed drinking water

Among paired households, 99.1% used an improved source, 85.5% on premises and 91.1% had sufficient water when needed (5,929 households that answered). All three criteria were met by 77.9% (76.6–79.2) of paired households (Fig 3). The JMP-aligned estimate was 42.5% (40.8– 44.1) of the household population, matching the official 42.6% (on-premises use 85.5% against 85.9%). Under a hypothetical scenario in which the source *E. coli* criterion was replaced with POU *E. coli* status, the estimated proportion meeting all safely managed criteria would decrease to 12.6% (11.4–13.8); this represents an illustrative monitoring scenario rather than a revised SDG indicator. Requiring both source and POU to be negative gave 10.6% (9.5–11.7). These are shares of the household population; the 12.7% ND/ND share above is a household share and a different quantity. Of the population meeting the JMP-aligned definition, 75.1% (72.6–77.4) had *E. coli* in the POU sample. The POU-substituted figure was 17.2% in urban and 10.6% in rural households, and 22.8% in the richest and 8.1–12.1% in the other quintiles. By division it ranged from 4.4% in Khulna and 5.3% in Barishal to 30.2% in Mymensingh, against JMP-aligned figures of 24.3% in Khulna to 63.0% in Rangpur; the gap was largest in Rangpur (63.0% to 14.1%) and Mymensingh (58.4% to 30.2%).

**Fig 3.**
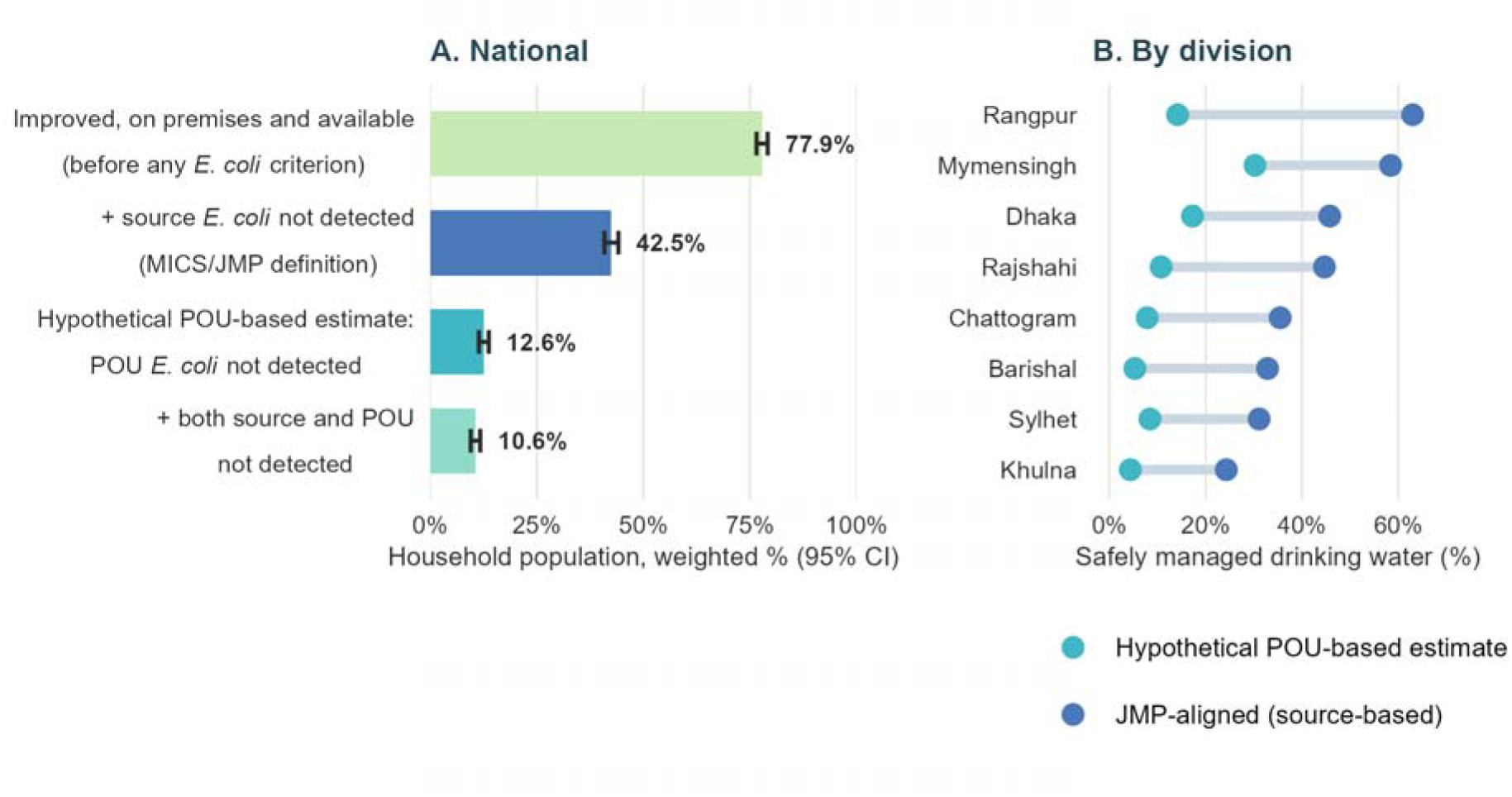
JMP-aligned estimate of safely managed drinking water and a hypothetical POU-based estimate in which the source *E. coli* criterion is replaced with POU *E. coli* status. (A) National estimates for the household population. (B) By division. The hypothetical POU-based estimate is not an alternative SDG 6.1 indicator; it illustrates how the estimated coverage would change if contamination were assessed at the point of use rather than at the source.

### Factors associated with POU detection where the source was negative

These analyses are secondary and describe associations, not causal effects. Among 2,933 complete-case households with a negative source (Table 4), glasses poured directly from the source were less often positive than those from covered containers (adjusted PR [aPR] 0.85, 0.77–0.94); uncovered and covered containers did not clearly differ after adjustment (1.05, 0.98– 1.13; unadjusted 1.07, 1.01–1.13), also among stored water only (1.05, 0.98–1.12). Estimates for no effective treatment (1.10, 0.95–1.27), no observed handwashing place with water and soap (1.04, 0.99–1.10) and a round trip of 30 minutes or more (1.10, 0.94–1.28; 31 households) were close to 1 with wide intervals. They were not consistent across models: among stored water only, the handwashing estimate was 1.06 (1.01–1.11), and in all paired households (Table 5) no effective treatment was 1.15 (1.06–1.26). Rural residence was associated with POU detection (1.11, 1.04–1.19). On the absolute scale, mutually adjusted prevalence differences were −11.1 percentage points (−17.3 to −4.9) for glasses poured directly from the source, +4.1 (−1.3 to 9.5) for uncovered containers, +5.7 (−3.3 to 14.7) for no effective treatment and +7.9 (3.2 to 12.6) for rural residence; in all paired households, a detected source was associated with +17.4 points (15.3 to 19.5) and no effective treatment with +10.6 (4.5 to 16.8) (S1 File). All four lower wealth quintiles had higher PRs than the richest (minimally adjusted 1.14–1.18), but with mutual adjustment only the second and fourth quintiles excluded 1 (1.12 each).

**Table 4.** Factors associated with POU *E. coli* detection among households with no detectable source *E. coli* (n = 2,933; modified Poisson, survey-weighted).

| Variable | Unweighted n | Weighted POU detection, % | Unadjusted PR (95% CI) | Minimally adjusted PR (95% CI)† | Mutually adjusted PR (95% CI)‡ |
| --- | --- | --- | --- | --- | --- |
| Container at POU test |  |  |  |  |  |
| Covered container (ref) | 1,896 | 76.9 | 1.00 | 1.00 | 1.00 |
| Direct from source | 422 | 62.5 | 0.81 (0.74–0.89) | 0.85 (0.78–0.94) | 0.85 (0.77–0.94) |
| Uncovered container | 615 | 82.3 | 1.07 (1.01–1.13) | 1.05 (0.98–1.13) | 1.05 (0.98–1.13) |
| Effective household treatment |  |  |  |  |  |
| Yes (ref) | 166 | 60.4 | 1.00 | 1.00 | 1.00 |
| No | 2,767 | 77.1 | 1.28 (1.11–1.47) | 1.08 (0.94–1.25) | 1.10 (0.95–1.27) |
| Handwashing place with water and soap |  |  |  |  |  |
| Observed (ref) | 2,047 | 73.6 | 1.00 | 1.00 | 1.00 |
| Not observed | 886 | 81.3 | 1.10 (1.05–1.16) | 1.04 (0.99–1.09) | 1.04 (0.99–1.10) |
| Collection round trip |  |  |  |  |  |
| On premises or <30 min (ref) | 2,902 | 75.7 | 1.00 | 1.00 | 1.00 |
| ≥30 min | 31 | 90.6 | 1.20 (1.04–1.38) | 1.09 (0.95–1.24) | 1.10 (0.94–1.28) |
| Wealth quintile |  |  |  |  |  |
| Richest (ref) | 511 | 63.0 | 1.00 | 1.00 | 1.00 |
| Fourth | 533 | 77.1 | 1.22 (1.12–1.34) | 1.15 (1.05–1.26) | 1.12 (1.02–1.23) |
| Middle | 560 | 77.8 | 1.24 (1.12–1.36) | 1.14 (1.03–1.26) | 1.09 (0.98–1.21) |
| Second | 610 | 81.8 | 1.30 (1.19–1.42) | 1.18 (1.08–1.29) | 1.12 (1.01–1.24) |
| Poorest | 719 | 80.7 | 1.28 (1.18–1.40) | 1.16 (1.06–1.28) | 1.09 (0.98–1.21) |
| Rural vs urban | 2,128 / 805 | 79.9 / 66.6 | 1.20 (1.12–1.28) | 1.13 (1.06–1.21) | 1.11 (1.04–1.19) |

*Unadjusted: single-exposure model. †Each row from a model adjusted for wealth, residence, division and head's education (exposure-specific; wealth and residence rows from the container model). ‡Model with all handling factors plus wealth, residence, division, head's education, source type and sanitation (S1 File). Source type is included because, although all households here already have a negative source result by construction, piped, deep-tubewell and shallow-tubewell supplies differ systematically in infrastructure and maintenance and may still capture residual heterogeneity in post-source contamination risk; it does not indicate a difference in source detection, which is fixed at zero by the negative-source restriction.*

**Table 5.**
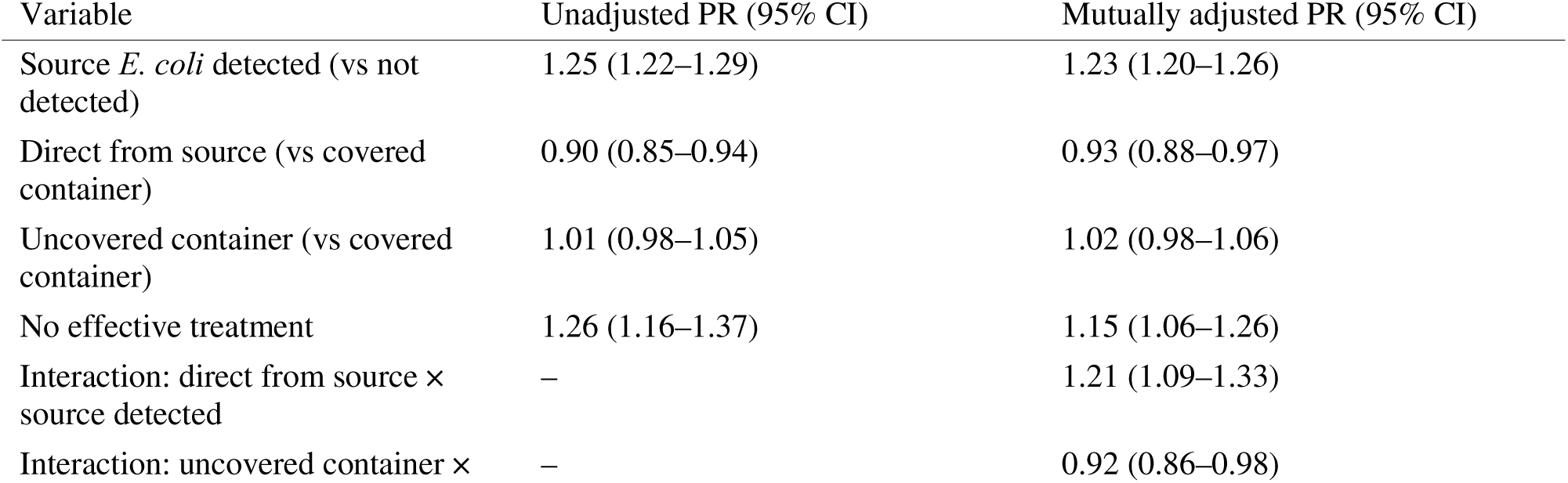
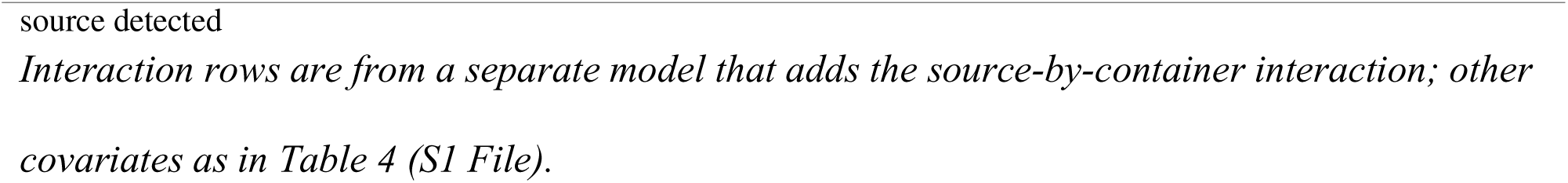
Factors associated with POU *E. coli* detection in all paired households, source status as a covariate (n = 5,714 complete cases; modified Poisson).

Results were similar in the 2,451 households whose glass source matched the main source (direct from source 0.85, 0.77–0.94; uncovered 1.04, 0.97–1.12; no effective treatment 1.10, 0.93–1.30), for on-premises sources (uncovered 1.05, 0.98–1.13), excluding households reporting treatment (1.04, 0.97–1.12) and with a month covariate (1.06, 0.99–1.13); POU detection was higher in June–July than February–March (1.15, 1.07–1.23). The container result depended on the threshold: at ≥11 CFU/100 mL uncovered versus covered containers gave 1.13 (1.04–1.24) and no effective treatment 1.21 (1.00–1.46) (S1 File). In all paired households (Table 5), a detected source was associated with POU detection (PR 1.23, 1.20–1.26) (94.7% versus 75.8% POU detection with and without source detection). Interaction estimates between container and source status are in Table 5 and S1 File. Pooling direct-from-source glasses with covered containers as the reference gave a PR of 1.09 (1.02–1.17) for uncovered containers.

### Divisional patterns

Among all households with POU *E. coli*, 47.0% (45.4–48.6) had a negative source. This share was highest in Rangpur (66.1%), Mymensingh (60.9%) and Barishal (58.5%) and lowest in Khulna (33.4%) and Sylhet (38.1%). Divisional samples are small (for example, n = 345 in Mymensingh). These estimates describe variation within this survey sample and are not rankings of divisional water quality.

### Childhood diarrhoea

Two-week diarrhoea prevalence was 6.1% (5.0–7.3) among 2,249 children. Compared with children in households without POU *E. coli*, those in households with POU detection had an adjusted PR of 1.04 (0.59–1.86; unadjusted 1.11, 0.62–1.98), and source detection alone gave 1.09 (0.75–1.58). In the exploratory four-pattern analysis (Table 6), prevalence was 5.5–6.3% across patterns and, relative to ND/ND, adjusted PRs were 1.02 (0.53–1.96) for ND/Detected, 1.20 (0.26–5.56) for Detected/ND and 1.10 (0.58–2.11) for Detected/Detected; adjusting only for age group and wealth gave 1.03, 1.23 and 1.11. In the same model, children in the poorest quintile had a higher prevalence than those in the richest (PR 2.30, 1.17–4.50), and children aged 36–47 and 48–59 months a lower prevalence than those under 12 months (0.36, 0.20–0.67; 0.38, 0.19–0.74) (S1 File). Only 14 children in the reference group had diarrhoea, and the smallest detectable PR was about 2.1–2.2, so moderate to large associations cannot be excluded. Results were the same with the under-five child weight (1.02, 0.53–1.96; 1.21, 0.26–5.64; 1.10, 0.57–2.10; n = 2,248) and a month covariate (1.05, 1.23, 1.11). Other sensitivity analyses gave imprecise estimates that varied in direction (S1 File). A ≥11 CFU/100 mL analysis gave a PR of 3.10 (1.32–7.31) for the rare Detected/ND pattern, based on very few children (S1 File).

**Table 6.**
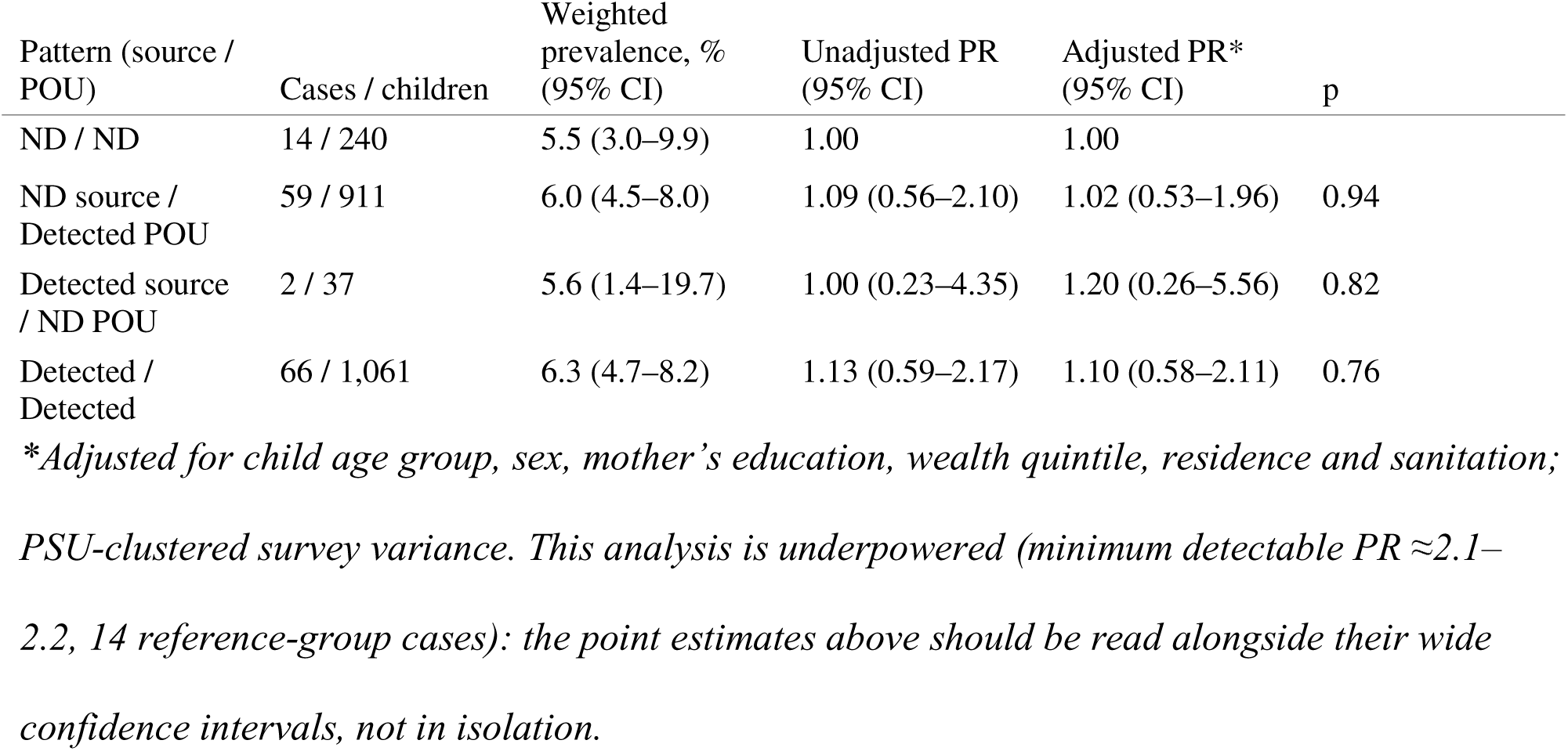
Joint source–POU status and two-week diarrhoea, children aged 0–59 months (n = 2,249; exploratory).

### Spatial clustering and geospatial covariates (exploratory)

POU detection was not randomly distributed across survey clusters. Global Moran’s I on the cluster-level detection rate (n = 3,145 GPS-matched clusters) was 0.160 (z = 16.5, p < 0.001), and the district-level test (n = 64 districts) gave 0.31 (z = 4.0, p < 0.001), indicating spatial clustering at both scales (Fig 4). District-level detection ranged from complete (100%) detection in five districts (Barguna, Jhalokati, Kushtia, Patuakhali and Sunamganj; 84–94 households tested in each, with no negative results) to 53–65% in the lowest-detection districts (S1 File); because a field-team or measurement-batch effect specific to these clusters cannot be ruled out (see below), this pattern should not be read as a ranking of district water safety. The exact 95% Clopper-Pearson CI for these five districts (95.7–100% to 96.2–100%) excludes the national POU rate (84.8%) by a wide margin (the probability of 84–94 consecutive detections under the national rate is between 2×10^−7^ and 9×10^−7^), so sampling chance under a uniform national rate does not explain these five districts; whether a non-sampling explanation specific to these districts (a genuinely higher local rate, or a systematic factor such as field-team assignment, discussed below) is responsible cannot be distinguished from these data alone.

**Fig 4.**
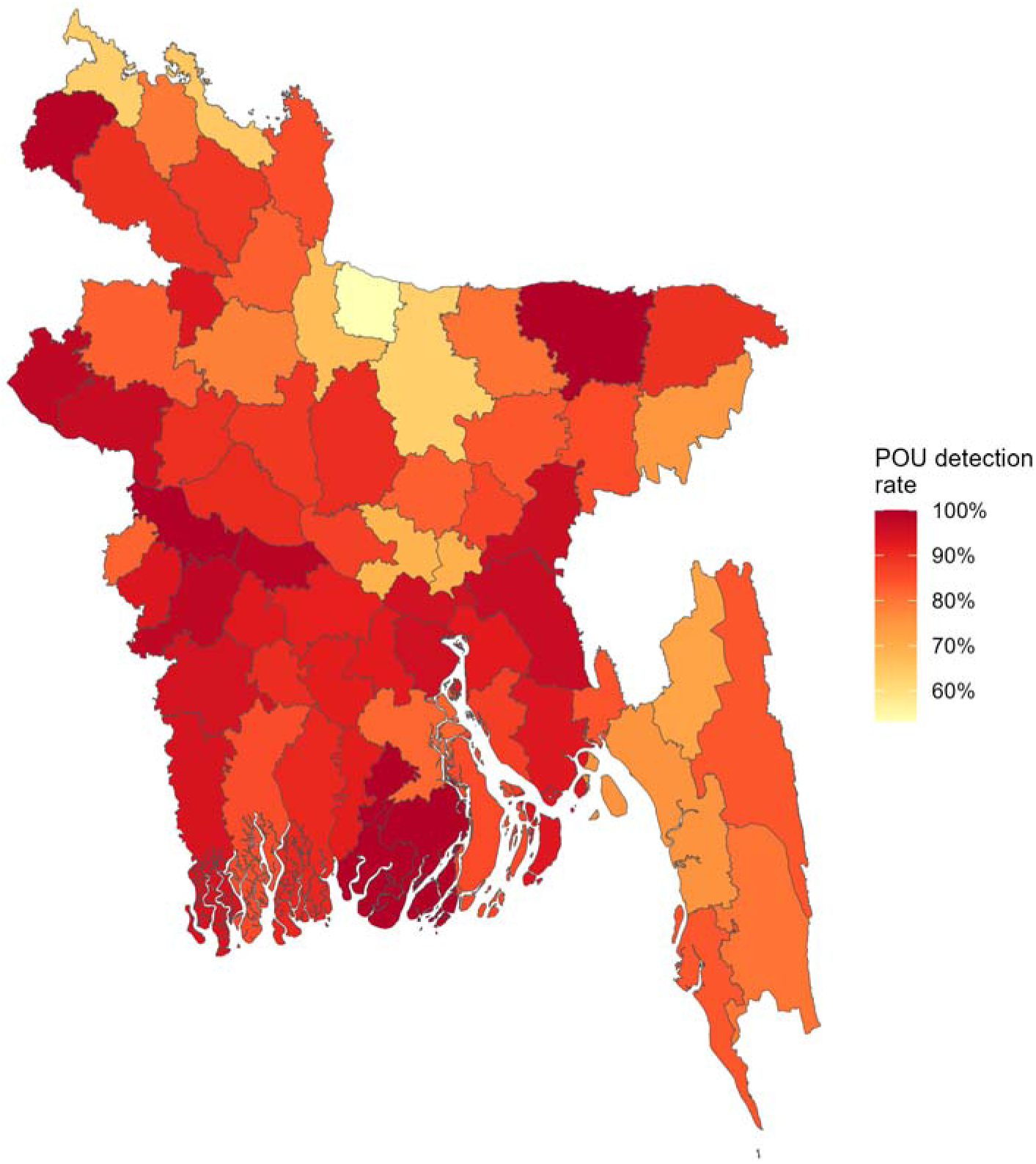
District-level POU *E. coli* detection, Bangladesh MICS 2025. Design-based district estimates (64 map units; city corporations are shown within their districts; administrative boundaries from an independent open geospatial database [28]). The colour scale spans the observed range (53% to 100%). The five darkest districts had complete detection; this is a signal for local verification, not a ranking of district water safety.

In a district-level ecological model (67 district-level domains: 64 districts and three city corporations), the five covariates jointly explained part of the variation between districts (F(5,61) = 5.67, p < 0.001; adjusted R^2^ = 0.26). Lower elevation (p = 0.014) and lower nightlights (p = 0.028) were independently associated with higher detection. Rainfall, population density and travel time were not (all p > 0.29). The result was similar without weights. It was weaker when districts with complete detection received inverse-variance weights (p = 0.048), because their zero standard error gives them very large weight (S1 File).

In the survey-weighted household-level mixed model (n = 6,048; 3,145 PSUs), lower elevation (OR 0.62 per SD, 95% CI 0.56–0.70, p<0.001) and longer travel time to healthcare (OR 1.65, 95% CI 1.29–2.12, p<0.001) were independently associated with higher odds of POU detection, and lower nightlights (a proxy for economic activity) was independently associated with higher odds (OR 0.82, 95% CI 0.70–0.97, p = 0.016); rainfall and population density were not independently significant, and the association with urban residence was attenuated to borderline significance once properly survey-weighted (OR 0.75, 95% CI 0.56–1.00, p = 0.050). Substantial between-PSU variance remained after adjustment (approximate intraclass correlation 0.47), indicating that the covariates available explain only part of the clustering.

## Discussion

### Principal findings

In a paired national sample, *E. coli* was detected in 47% of source samples and 85% of POU samples, and three in four households with a negative source had *E. coli* in the glass of water served. Only about one in eight households was negative at both locations. Substituting POU for source *E. coli* in the safely managed calculation lowered the estimate from 42.5% to 12.6%; this is a counterfactual calculation, not an alternative estimate of the SDG indicator. Discordance persisted when the glass was poured directly from the source. No handling factor was consistently associated with POU detection apart from glasses poured directly from the source, rural residence and (less consistently) lower wealth; POU detection also clustered geographically; and estimates of the association with diarrhoea were imprecise.

### Source and point-of-use detection, and their discordance

The source sample is taken by field staff at the source; the POU sample is a glass of the water the household usually drinks, which may have been collected earlier and may not come from the sampled source. The contrast therefore combines biological heterogeneity, temporal variability, differences between the two sampling locations and procedural differences. Random measurement error around a shared concentration would produce discordance in both directions; the 16:1 imbalance therefore reflects a systematic difference between the two samples, but the data cannot say whether it is post-collection contamination or a feature of the sampling. The direct-from-source benchmark points towards the second: discordance persisted with almost no storage time (33.3% ND/Detected, against 39.0% and 48.3% for covered and uncovered containers), so storage explains only part of it. A related concern is that in 17% of households the recorded glass source differed from the main source tested, so some apparent discordance could reflect households drinking from a different, untested source rather than post-collection change in the same water; restricting to the 4,963 households whose glass source matched the main source left the discordance essentially unchanged (ND/Detected 39.8% versus 39.9% in the full sample), which argues against source-switching as a major driver of the overall pattern, though it cannot rule out switching within specific households. The rare Detected/ND pattern (2.5%) could reflect household treatment, intermittent source contamination or flushing, which we did not examine. The mineral-water blanks (1.2% positive) test the filtration and laboratory steps and cannot rule out contamination through the glass, hands or pouring. Washed drinking vessels in low-income Dhaka households carried a mean of 167 *E. coli* CFU per 10 cm^2^ [29], so the glass itself is a plausible route. Such contamination would still be real exposure, because household members drink from these vessels.

The findings agree with paired studies elsewhere [9,10], with meta-analytic evidence that stored water is more often contaminated than source water [12], and with the multi-country MICS analysis in which POU contamination was more frequent than source contamination and higher in rural and wet-season samples [15]. The rise in detection from February–March to June–July is in the same direction as that multi-country pattern. Bangladesh’s 47.4% source and 84.8% POU detection fall within the ranges across those 27 surveys [15]. Those ranges were 16% to 90% at the point of collection and 19% to 99% at the POU. In a low-income Dhaka community, same-day pairs showed *E. coli* in 58% of source and 77% of drinking-vessel samples [30]. There, a clean source with contaminated drinking water outnumbered the reverse by about 1.8 to 1 (314 versus 177 pairs) [30]. The 16:1 imbalance in this national sample is far larger. Differences in setting, sampling and laboratory method may explain part of the gap.

### Implications for safely managed water monitoring

The source-based safely managed indicator is defined at the source, and we do not propose changing it. In one national sample the source and household classifications differed frequently, and the estimate changes substantially if POU *E. coli* is substituted. MICS already tests both locations [8,15], so the question is whether POU measurement adds useful information. The POU-substituted figure is an illustrative diagnostic of post-source contamination, not a revised indicator: source quality reflects the performance of water supply services, whereas POU quality also reflects household storage, handling and hygiene, and the two carry different accountability, though this design cannot establish whether the discordance reflects contamination after the source measurement or differences in sampling and handling between locations. Source and POU classifications clearly differ in this national sample, but that does not mean the JMP indicator itself is wrong: we did not test the indicator, and we cannot show whether POU testing would predict enteric illness better than source testing or whether routine POU testing is justified globally. What the data can support is a narrower claim, that POU measurement could add information as a complement to source testing.

If the difference mainly reflects contamination after collection, a POU measure would give an indicator of microbiological contamination closer to the point of consumption (*E. coli* indicates faecal contamination and is not a direct measure of pathogen exposure). If it mainly reflects the sampling procedures, the priority is to standardise the POU sample, for example by also sampling stored water in a sterile container and testing source and POU on the same visit with a recorded time gap. Before any change to monitoring, future work should test whether the difference is reproducible across seasons and countries, how a standardised POU sample performs, and whether POU results add predictive value for enteric disease beyond source results, at what cost and with what implications for comparability across countries and years. Bangladesh is one national case, so these findings call for multi-country validation rather than modification of the global indicator.

### Factors associated with point-of-use detection

Apart from glasses poured directly from the source, only rural residence and, less consistently, lower wealth were associated with POU detection (Table 4); container type, treatment and handwashing showed no consistent association. This pattern is in the same direction as prior work. In rural Bangladesh, POU *E. coli* among deep-tubewell users rose with source contamination and walking time to the source [31]; in our data a detected source was associated with POU detection, and long collection trips were too few (31 households) to add to this. Treatment arms in the WASH Benefits trial roughly halved *E. coli* prevalence in stored water [32]. However, self-reported treatment in surveys tracks protection poorly: in 25 countries many households reporting treatment still had contaminated stored water [33], as in ours (60% POU detection among negative-source households reporting effective treatment). In a Dhaka community, 78% of treated and 76% of untreated drinking-water samples contained *E. coli* [30]. Our treatment estimates cannot be read as effectiveness, because households may treat water precisely because they perceive it as contaminated.

The container results should not be turned into storage advice. Glasses poured directly from the source were less often positive, which is expected with little storage time. The uncovered-container difference appeared only at the ≥11 CFU/100 mL threshold, a comparison we did not prespecify, and pooling direct-from-source glasses with covered containers would create a modest positive association (1.09, 1.02–1.17) that reflects the composition of the reference group. The share of POU-positive households with a negative source ranged from 33% in Khulna to 66% in Rangpur. Divisional samples are small and cross-sectional, so we do not rank divisions; programme decisions would need local investigation. Trial evidence supports household water treatment (filtration risk ratio 0.50 and chlorination 0.66 for diarrhoea in a recent meta-analysis [34]), but the large Bangladesh trial found no clear effect of water treatment alone [35]. Controlled studies of storage and serving practices are warranted. Because handling factors were weakly and inconsistently associated with POU detection and prevalence was high in nearly every group, household handling interventions alone may not fully address contamination when source and supply-chain factors contribute; source protection and safe management of the whole supply chain should be considered alongside them.

### Childhood diarrhoea

The prevalence ratio for POU detection and two-week diarrhoea was close to 1 but too imprecise to interpret (1.04, 0.59–1.86), because the water-quality subsample contains two households per PSU and only 14 reference children had diarrhoea. This estimate is compatible with both no effect and a moderate to large one, so it neither confirms nor rules out a water-quality contribution to diarrhoea in this sample. Analyses of MICS 2019 reported higher odds of diarrhoea in children from households with moderate or high POU *E. coli* [17]; comparison with that earlier analysis is limited by our imprecision, not by disagreement in direction. A prospective cohort in Dhaka slums (CHoBI7 programme) linked high source contamination to diarrhoea (odds ratio 1.43 for source *E. coli* >100 CFU/100 mL) [36]. In the same model, wealth was associated with diarrhoea far more strongly than any water-quality measure, consistent with structural determinants shaping child health more than household water handling. Two-week caregiver-reported diarrhoea after one water sample cannot exclude reverse causation or non-water routes. For context, a single-date *E. coli* count did not predict bacterial pathogens in Bangladesh tubewells [37], and environmental pathogen detection in a pooled analysis was associated with child infection but not with diarrhoea overall [38]. A null result here should not be read as absence of an effect.

### Geographic clustering

POU detection clustered geographically rather than being randomly scattered across clusters and districts (Moran’s I 0.16–0.31, p<0.001 at both scales), and this clustering was only partly explained by elevation, travel time to healthcare and the other geospatial covariates available. Independent, unbiased household-level measurement error around a shared national concentration would not, by itself, generate spatial autocorrelation of this magnitude or produce the five districts’ exact confidence intervals reported above, which exclude the national rate by a wide margin; the finding is therefore inconsistent with pure random sampling noise. This is a narrower claim than saying the pattern reflects a real environmental exposure: MICS fieldwork is organised so that survey teams and equipment batches cover contiguous PSUs, so a geographically clustered field-team or batch effect (for example, a team with different pouring or handling technique, or a defective lot of filtration equipment) would also produce spatial autocorrelation and is not distinguishable from a true exposure gradient using the data reported here, since measurer or team identifiers were not linked to the spatial analysis. We therefore treat the geographic pattern as a real, non-random signal requiring further investigation, not as established evidence of an environmental exposure gradient.

This distinction matters for how the finding should be used. Flagging the five complete-detection districts (84–94 households each) for local investigation is a reasonable next step precisely because the pattern is unlikely to be sampling noise, whatever its cause. Whether the same GPS-linked geospatial covariates could be produced routinely for every MICS round at low marginal cost is plausible in principle, since the underlying raster layers are already compiled by the MICS GIS programme, but this analysis does not establish the analytic capacity, review burden or cost of institutionalising it, and we do not make that claim. Coordinates are displaced within the local administrative unit and the covariates are ecological, so this spatial analysis identifies areas for investigation, not individual household risk (Strengths and limitations).

### Strengths and limitations

Strengths are national representation, standardised testing, paired samples from the same households, design-based inference and validation against the official indicators. Five limitations matter most. First, the design is cross-sectional: both samples come from one visit, so the sequence of contamination and causal effects cannot be established; treatment and storage may respond to perceived water quality, and the models are descriptive. Second, the samples are not directly comparable: the two differ in location and procedure, flushing, storage time, glass cleanliness and the time elapsed between the fixed household-then-source collections are undocumented, the glass source differed from the main source in 17% of households, and the POU sample is a glass, not necessarily stored water. Third, selection may bias the estimates: households lost from the paired sample were more often off premises (38% versus 15%) or used other sources (23% versus 2%), and 126 of 2,377 children (5.3%) lacked an under-five questionnaire; weighting corrects only for observed characteristics, so the estimates may not represent these groups. Fourth, precision is limited: two households per PSU limit subgroup precision, only 14 reference children had diarrhoea (smallest detectable PR about 2.1–2.2), ratio measures are constrained when prevalence exceeds 90%, and the 6.1% diarrhoea prevalence is not a national under-five estimate. Diarrhoea recall timing relative to water sampling, and possible correlated illness among siblings, could not be verified or adjusted for beyond PSU-level clustering. Fifth, scope and generalisability are limited in several ways. The spatial analysis was exploratory and not prespecified; MICS displaces cluster coordinates within the local administrative unit for confidentiality, so results describe broad spatial patterns rather than precise locations; the geospatial covariates are ecological (cluster- or district-level averages, subject to the modifiable areal unit problem), so associations with them cannot be interpreted as individual-level exposures; and although confidence intervals for the five complete-detection districts exclude the national rate (S1 File), a field-team or measurement-batch effect cannot be ruled out, so these districts should be treated as candidates for local verification rather than an official ranking of water safety. This analysis also addresses microbiological safety only: MICS 2025 separately tested and reported source-water arsenic (Tables WS.1.6S2, WS.1.8), a well-documented chemical safety burden in Bangladesh groundwater [39] that the JMP safely managed definition also requires freedom from, but this analysis does not incorporate it, so source-type associations reported here should not be read as an overall safety ranking. Finally, time and place limit generalisability: testing took place from February to June in one country, before the monsoon peak [40], and POU detection increased across this window, so the study period may not capture the full range of seasonal variation, particularly the monsoon months when contamination is often highest; the POU-substituted figure is a hypothetical estimate for the survey period, and the safely managed calculation uses the *E. coli* criterion only, as MICS does.

### Conclusions

In the Bangladesh MICS 2025 water-quality subsample, source and household point-of-use *E. coli* classifications differed substantially within households. Replacing the source criterion with POU *E. coli* gave a substantially lower hypothetical monitoring estimate of safely managed drinking water (12.6% versus 42.5%), but this comparison cannot establish when or where contamination occurred. Among households with no detectable *E. coli* in the source sample, *E. coli* was frequently detected in the household drinking-water sample, including when the glass was poured directly from the source, and no handling factor was consistently associated with POU detection apart from this, rural residence and, less consistently, lower wealth. The estimates for diarrhoea were too imprecise to draw conclusions about health. POU contamination clustered geographically rather than appearing randomly distributed, and this clustering was only partly explained by elevation, travel time to healthcare and other geospatial covariates, with five districts showing complete detection in this sample. The findings support evaluating standardised POU sampling as a complement to source-based monitoring, including reproducibility across seasons and countries, incremental predictive value, feasibility and cost, and support geographically targeted follow-up investigation in the highest-detection districts using data Bangladesh already collects.

## Data Availability

All data analyzed in this study are available online from the UNICEF MICS repository at https://mics.unicef.org/surveys (Bangladesh Multiple Indicator Cluster Survey 2025). Datasets are accessible to registered users upon application.

## Author contributions

SMAUH conceived the study, developed the methodology, curated the data, carried out the formal analysis, prepared the software and figures, and wrote the original draft. KUB supervised the study. MKH administered the project and the journal submission. MKI, MKH, MS, BB, ZZH and KUB critically reviewed the results and their interpretation. All authors reviewed and edited the manuscript and approved the final version.

## Acknowledgments

We thank the Bangladesh Bureau of Statistics and UNICEF for making the MICS 2025 data available, and the UNICEF MICS GIS programme for the GPS and geospatial covariate datasets. We also thank the field teams, water-quality measurers and households who took part in the survey.

## Supporting information

**S1 Fig.**
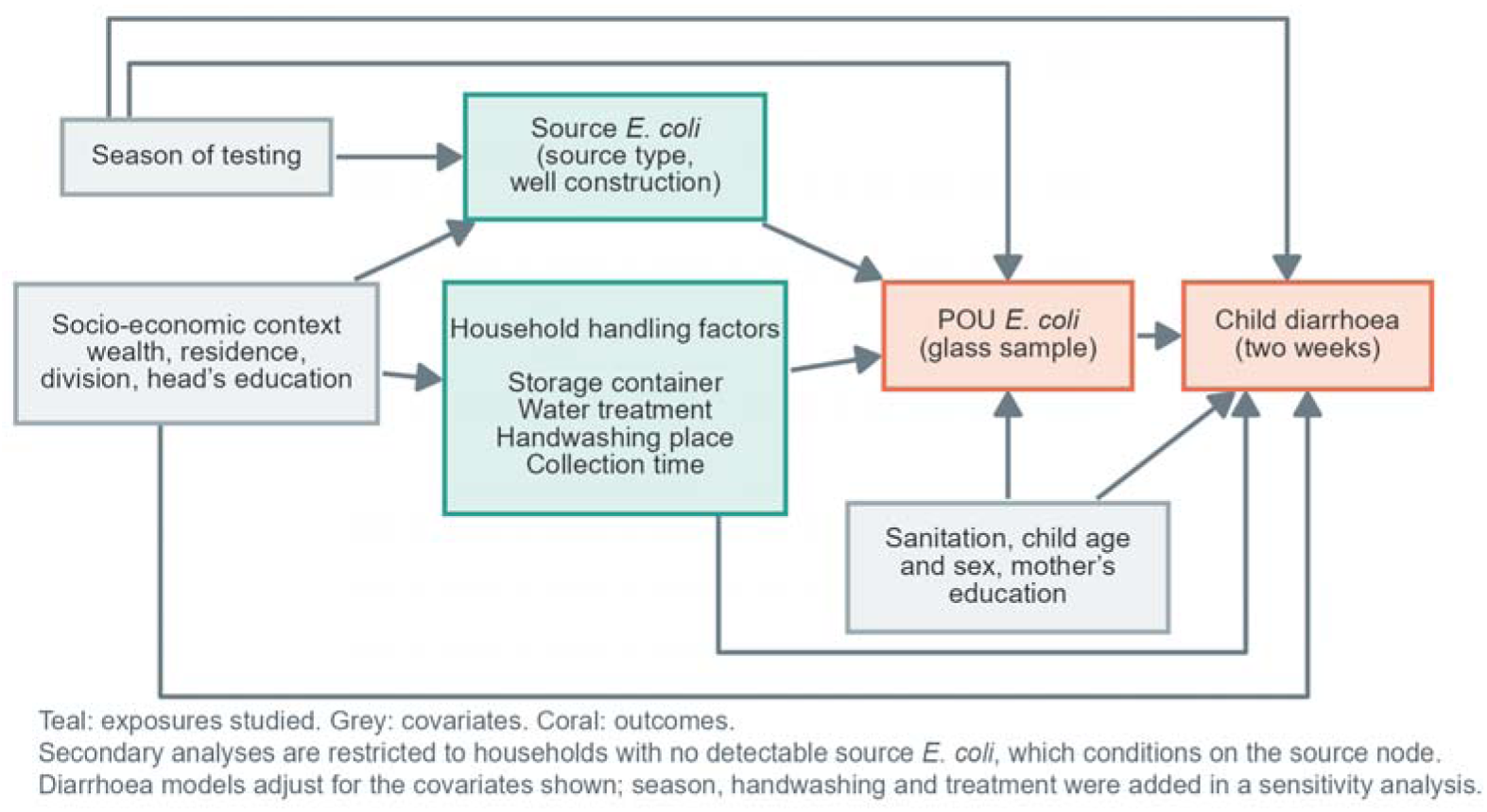
Conceptual framework (directed acyclic graph) for the analyses.

**S2 Fig.**
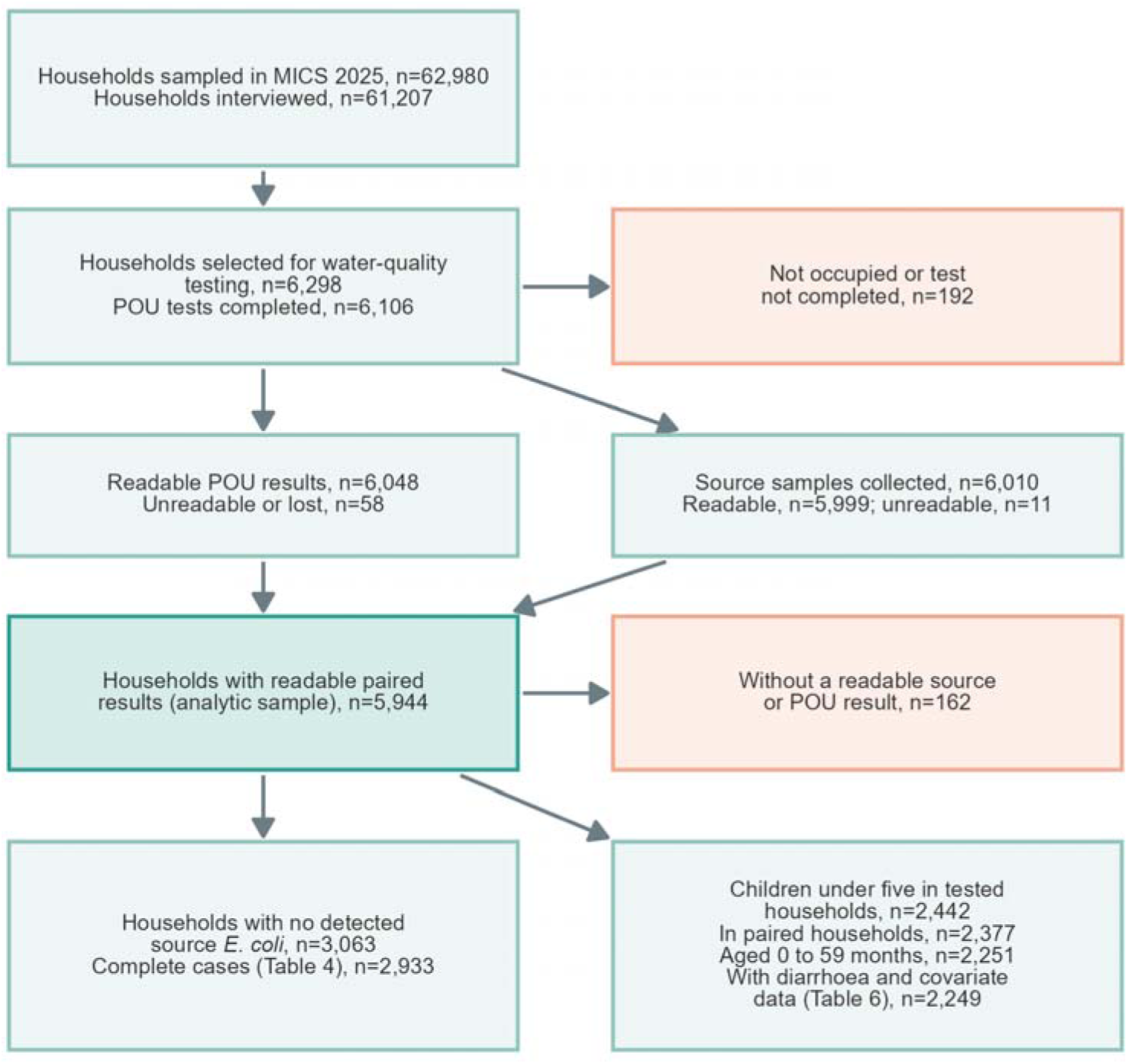
Flow of households and children into the analyses.

**S1 STROBE Checklist.** STROBE checklist for cross-sectional studies.

**S1 File.** Regression output for the main models (log prevalence ratios, standard errors, prevalence ratios with 95% confidence intervals and exact p-values), adjusted prevalence differences, summary tables of the sensitivity analyses, joint patterns at the ≥11 CFU/100 mL threshold, discordance by container and source type, the inverse-probability-weighting and glass-source sensitivity analyses, minimum detectable prevalence ratios, and the full district-level spatial regression and mixed-model output.

**S2 File.** Sample and missing-data audit: numbers at each step, missing values by variable, testing dates, blank controls, and included versus excluded households.

