## Supplementary material for "Discordance between source and point-of-use drinking-water *Escherichia coli* contamination in Bangladesh: a cross-sectional analysis and implications for safely managed water monitoring": S1_File_regression_output.docx

### S1 File. Regression output and sensitivity analyses for Tables 4–6

Survey-weighted modified Poisson regression (Stata svy: poisson, log link, linearised SE) gives prevalence ratios (PR); adjusted prevalence differences come from survey-weighted linear probability models (svy: regress) and are in percentage points. Full model output is given for the main models (Tables 4, 5 and 6); sensitivity analyses are summarised in single tables in which the PR for each exposure is shown across specifications (covariates other than the exposure of interest are those of the corresponding model and their estimates were stable). Survey Poisson models are fitted by pseudolikelihood, so likelihood-based goodness-of-fit statistics are not available; the number of observations, design degrees of freedom and PSUs are given for each full model. Terms are labelled by category rather than by their Stata variable code; the reference category for each factor is given in parentheses on the first row of that factor (for example “Poorest (vs richest)” is the wealth-quintile term, with the richest quintile as reference).

#### Table 4, mutually adjusted model (n = 2,933; design df = 3,018; PSUs in design = 3,149)

| Term | log(PR) | SE | PR (95% CI) | p |
| --- | --- | --- | --- | --- |
| Direct from source (vs covered container) | -0.161 | 0.049 | 0.85 (0.77–0.94) | <0.001 |
| Uncovered container (vs covered container) | 0.052 | 0.035 | 1.05 (0.98–1.13) | 0.132 |
| No effective treatment | 0.093 | 0.073 | 1.10 (0.95–1.27) | 0.203 |
| No handwashing place with water and soap | 0.042 | 0.025 | 1.04 (0.99–1.10) | 0.097 |
| Round trip ≥30 min | 0.093 | 0.079 | 1.10 (0.94–1.28) | 0.243 |
| Poorest (vs richest) | 0.086 | 0.055 | 1.09 (0.98–1.21) | 0.119 |
| Second (vs richest) | 0.112 | 0.051 | 1.12 (1.01–1.24) | 0.029 |
| Middle (vs richest) | 0.083 | 0.053 | 1.09 (0.98–1.21) | 0.118 |
| Fourth (vs richest) | 0.110 | 0.048 | 1.12 (1.02–1.23) | 0.021 |
| Rural (vs urban) | 0.108 | 0.034 | 1.11 (1.04–1.19) | 0.001 |
| Barishal (vs Dhaka) | 0.124 | 0.043 | 1.13 (1.04–1.23) | 0.004 |
| Chattogram (vs Dhaka) | 0.082 | 0.042 | 1.09 (1.00–1.18) | 0.050 |
| Khulna (vs Dhaka) | 0.108 | 0.042 | 1.11 (1.03–1.21) | 0.011 |
| Mymensingh (vs Dhaka) | -0.266 | 0.082 | 0.77 (0.65–0.90) | 0.001 |
| Rajshahi (vs Dhaka) | 0.053 | 0.046 | 1.05 (0.96–1.15) | 0.251 |
| Rangpur (vs Dhaka) | -0.002 | 0.048 | 1.00 (0.91–1.10) | 0.974 |
| Sylhet (vs Dhaka) | 0.049 | 0.054 | 1.05 (0.95–1.17) | 0.361 |
| Head’s education: primary (vs none/pre-primary) | -0.031 | 0.030 | 0.97 (0.91–1.03) | 0.305 |
| Head’s education: secondary (vs none/pre-primary) | -0.009 | 0.029 | 0.99 (0.94–1.05) | 0.755 |
| Head’s education: higher secondary+ (vs none/pre-primary) | -0.113 | 0.047 | 0.89 (0.81–0.98) | 0.017 |
| Deep tube well (vs piped) | 0.061 | 0.044 | 1.06 (0.97–1.16) | 0.171 |
| Shallow tube well (vs piped) | 0.039 | 0.042 | 1.04 (0.96–1.13) | 0.357 |
| Other source (vs piped) | -0.096 | 0.123 | 0.91 (0.71–1.16) | 0.435 |
| Unimproved sanitation (vs improved) | 0.013 | 0.041 | 1.01 (0.93–1.10) | 0.750 |

#### Sensitivity of the Table 4 estimates across specifications (PR, 95% CI)

Minimally adjusted = wealth, residence, division and head’s education, one exposure at a time; mutually adjusted = all handling factors, source type and sanitation. Blank cells (–) indicate that the exposure was not estimable or not in the model (for example, collection time in on-premises households).

| Exposure | Unadjusted | Minimally adjusted | Mutually adjusted | Stored water only | On-premises source | Excluding treatment | Month covariate | Threshold ≥11 CFU/100 mL | Glass source matches main |
| --- | --- | --- | --- | --- | --- | --- | --- | --- | --- |
| Direct from source (vs covered) | 0.81 (0.74–0.89) | 0.85 (0.78–0.94) | 0.85 (0.77–0.94) | – | 0.84 (0.76–0.93) | 0.84 (0.76–0.92) | 0.85 (0.77–0.93) | 0.84 (0.76–0.94) | 0.85 (0.77–0.94) |
| Uncovered container (vs covered) | 1.07 (1.01–1.13) | 1.05 (0.98–1.13) | 1.05 (0.98–1.13) | 1.05 (0.98–1.12) | 1.05 (0.98–1.13) | 1.04 (0.97–1.12) | 1.06 (0.99–1.13) | 1.13 (1.04–1.24) | 1.04 (0.97–1.12) |
| No effective treatment | 1.28 (1.11–1.47) | 1.08 (0.94–1.25) | 1.10 (0.95–1.27) | 1.11 (0.95–1.29) | 1.11 (0.96–1.29) | – | 1.09 (0.95–1.26) | 1.21 (1.00–1.46) | 1.10 (0.93–1.30) |
| No handwashing place with water and soap | 1.10 (1.05–1.16) | 1.04 (0.99–1.09) | 1.04 (0.99–1.10) | 1.06 (1.01–1.11) | 1.06 (1.00–1.12) | 1.04 (0.99–1.09) | 1.05 (1.00–1.10) | 1.05 (0.98–1.12) | 1.03 (0.98–1.09) |
| Round trip ≥30 min | 1.20 (1.04–1.38) | 1.09 (0.95–1.24) | 1.10 (0.94–1.28) | 1.06 (0.91–1.24) | – | 1.09 (0.93–1.28) | 1.10 (0.94–1.29) | 1.07 (0.87–1.31) | 1.02 (0.84–1.25) |
| n (households) | 2,933 | 2,933 | 2,933 | 2,511 | 2,505 | 2,747 | 2,933 | 4,549 | 2,451 |

#### Table 5, all paired households, source status as covariate (n = 5,714; design df = 3,018; PSUs in design = 3,149)

| Term | log(PR) | SE | PR (95% CI) | p |
| --- | --- | --- | --- | --- |
| Source E. coli detected (vs not detected) | 0.205 | 0.013 | 1.23 (1.20–1.26) | <0.001 |
| Direct from source (vs covered container) | -0.075 | 0.024 | 0.93 (0.88–0.97) | 0.002 |
| Uncovered container (vs covered container) | 0.016 | 0.020 | 1.02 (0.98–1.06) | 0.428 |
| No effective treatment | 0.143 | 0.044 | 1.15 (1.06–1.26) | 0.001 |
| No handwashing place with water and soap | 0.014 | 0.013 | 1.01 (0.99–1.04) | 0.288 |
| Round trip ≥30 min | 0.045 | 0.031 | 1.05 (0.98–1.11) | 0.154 |
| Poorest (vs richest) | 0.071 | 0.028 | 1.07 (1.02–1.13) | 0.011 |
| Second (vs richest) | 0.092 | 0.026 | 1.10 (1.04–1.15) | <0.001 |
| Middle (vs richest) | 0.064 | 0.027 | 1.07 (1.01–1.12) | 0.017 |
| Fourth (vs richest) | 0.068 | 0.026 | 1.07 (1.02–1.13) | 0.010 |
| Rural (vs urban) | 0.047 | 0.018 | 1.05 (1.01–1.09) | 0.008 |
| Barishal (vs Dhaka) | 0.074 | 0.022 | 1.08 (1.03–1.13) | <0.001 |
| Chattogram (vs Dhaka) | 0.053 | 0.020 | 1.05 (1.01–1.10) | 0.010 |
| Khulna (vs Dhaka) | 0.048 | 0.019 | 1.05 (1.01–1.09) | 0.012 |
| Mymensingh (vs Dhaka) | -0.210 | 0.052 | 0.81 (0.73–0.90) | <0.001 |
| Rajshahi (vs Dhaka) | 0.029 | 0.022 | 1.03 (0.99–1.08) | 0.191 |
| Rangpur (vs Dhaka) | -0.003 | 0.027 | 1.00 (0.95–1.05) | 0.904 |
| Sylhet (vs Dhaka) | 0.021 | 0.026 | 1.02 (0.97–1.08) | 0.435 |
| Head’s education: primary (vs none/pre-primary) | -0.024 | 0.017 | 0.98 (0.94–1.01) | 0.142 |
| Head’s education: secondary (vs none/pre-primary) | -0.003 | 0.016 | 1.00 (0.97–1.03) | 0.825 |
| Head’s education: higher secondary+ (vs none/pre-primary) | -0.060 | 0.024 | 0.94 (0.90–0.99) | 0.014 |
| Deep tube well (vs piped) | 0.047 | 0.022 | 1.05 (1.00–1.10) | 0.035 |
| Shallow tube well (vs piped) | 0.027 | 0.021 | 1.03 (0.99–1.07) | 0.204 |
| Other source (vs piped) | -0.031 | 0.046 | 0.97 (0.89–1.06) | 0.506 |
| Unimproved sanitation (vs improved) | -0.011 | 0.021 | 0.99 (0.95–1.03) | 0.612 |

#### Table 5, all paired households, source status x container interaction (n = 5,714; design df = 3,018; PSUs in design = 3,149)

| Term | log(PR) | SE | PR (95% CI) | p |
| --- | --- | --- | --- | --- |
| Source E. coli detected (vs not detected) | 0.194 | 0.016 | 1.21 (1.18–1.25) | <0.001 |
| Direct from source (vs covered container) | -0.177 | 0.048 | 0.84 (0.76–0.92) | <0.001 |
| Uncovered container (vs covered container) | 0.057 | 0.031 | 1.06 (1.00–1.12) | 0.065 |
| Interaction: source detected × direct from source | 0.188 | 0.050 | 1.21 (1.09–1.33) | <0.001 |
| Interaction: source detected × uncovered container | -0.082 | 0.032 | 0.92 (0.86–0.98) | 0.012 |
| No effective treatment | 0.146 | 0.044 | 1.16 (1.06–1.26) | <0.001 |
| No handwashing place with water and soap | 0.012 | 0.013 | 1.01 (0.99–1.04) | 0.354 |
| Round trip ≥30 min | 0.044 | 0.032 | 1.04 (0.98–1.11) | 0.164 |
| Poorest (vs richest) | 0.070 | 0.028 | 1.07 (1.02–1.13) | 0.013 |
| Second (vs richest) | 0.092 | 0.026 | 1.10 (1.04–1.15) | <0.001 |
| Middle (vs richest) | 0.065 | 0.027 | 1.07 (1.01–1.12) | 0.016 |
| Fourth (vs richest) | 0.067 | 0.026 | 1.07 (1.01–1.13) | 0.012 |
| Rural (vs urban) | 0.049 | 0.018 | 1.05 (1.01–1.09) | 0.006 |
| Barishal (vs Dhaka) | 0.075 | 0.022 | 1.08 (1.03–1.13) | <0.001 |
| Chattogram (vs Dhaka) | 0.053 | 0.021 | 1.05 (1.01–1.10) | 0.010 |
| Khulna (vs Dhaka) | 0.054 | 0.019 | 1.06 (1.02–1.09) | 0.004 |
| Mymensingh (vs Dhaka) | -0.205 | 0.052 | 0.81 (0.74–0.90) | <0.001 |
| Rajshahi (vs Dhaka) | 0.026 | 0.022 | 1.03 (0.98–1.07) | 0.241 |
| Rangpur (vs Dhaka) | -0.013 | 0.027 | 0.99 (0.94–1.04) | 0.622 |
| Sylhet (vs Dhaka) | 0.017 | 0.026 | 1.02 (0.97–1.07) | 0.511 |
| Head’s education: primary (vs none/pre-primary) | -0.025 | 0.016 | 0.98 (0.94–1.01) | 0.132 |
| Head’s education: secondary (vs none/pre-primary) | -0.006 | 0.016 | 0.99 (0.96–1.03) | 0.698 |
| Head’s education: higher secondary+ (vs none/pre-primary) | -0.062 | 0.024 | 0.94 (0.90–0.99) | 0.011 |
| Deep tube well (vs piped) | 0.043 | 0.022 | 1.04 (1.00–1.09) | 0.054 |
| Shallow tube well (vs piped) | 0.023 | 0.021 | 1.02 (0.98–1.07) | 0.273 |
| Other source (vs piped) | -0.033 | 0.046 | 0.97 (0.88–1.06) | 0.475 |
| Unimproved sanitation (vs improved) | -0.010 | 0.021 | 0.99 (0.95–1.03) | 0.631 |

#### Adjusted prevalence differences (percentage points, 95% CI)

From survey-weighted linear probability models with the covariates of the corresponding PR model.

| Exposure | Table 4, mutually adjusted (n = 2,933) | Table 4, minimally adjusted | Table 5 (all paired households, n = 5,714) |
| --- | --- | --- | --- |
| Direct from source (vs covered container) | -11.1 (-17.3 to -4.9); p = <0.001 | -10.9 (-17.1 to -4.7); p = <0.001 | -6.1 (-9.8 to -2.4); p = 0.001 |
| Uncovered container (vs covered container) | +4.1 (-1.3 to +9.5); p = 0.134 | +4.0 (-1.3 to +9.4); p = 0.137 | +1.3 (-2.0 to +4.7); p = 0.432 |
| No effective treatment | +5.7 (-3.3 to +14.7); p = 0.217 | +4.8 (-4.1 to +13.7); p = 0.293 | +10.6 (+4.5 to +16.8); p = <0.001 |
| No handwashing place with water and soap | +3.3 (-0.7 to +7.2); p = 0.105 | +3.1 (-0.9 to +7.1); p = 0.128 | +1.2 (-1.0 to +3.4); p = 0.288 |
| Round trip ≥30 min | +8.4 (-5.3 to +22.1); p = 0.230 | +7.5 (-4.6 to +19.6); p = 0.223 | +4.4 (-1.4 to +10.2); p = 0.138 |
| Rural (vs urban) | +7.9 (+3.2 to +12.6); p = 0.001 | +8.9 (+4.2 to +13.5); p = <0.001 | +3.9 (+1.1 to +6.8); p = 0.007 |
| Source E. coli detected (vs not detected) | – | – | +17.4 (+15.3 to +19.5); p = <0.001 |

#### Table 5 model in households whose glass source matches the main source

| Table 5 term (glass source matches main source; n = 4,773) | PR (95% CI) |
| --- | --- |
| Source E. coli detected | 1.22 (1.19–1.26) |
| Direct from source | 0.92 (0.88–0.97) |
| Uncovered container | 1.01 (0.97–1.05) |
| No effective treatment | 1.15 (1.04–1.27) |
| No handwashing place | 1.02 (0.99–1.04) |
| Round trip ≥30 min | 1.01 (0.94–1.09) |

#### Table 6, diarrhoea, main model (n = 2,249; design df = 1,596; PSUs in design = 1,727)

| Term | log(PR) | SE | PR (95% CI) | p |
| --- | --- | --- | --- | --- |
| ND source / Detected POU (vs ND/ND) | 0.023 | 0.331 | 1.02 (0.53–1.96) | 0.945 |
| Detected source / ND POU (vs ND/ND) | 0.180 | 0.782 | 1.20 (0.26–5.56) | 0.818 |
| Detected / Detected (vs ND/ND) | 0.099 | 0.329 | 1.10 (0.58–2.11) | 0.764 |
| Age 12–23 months (vs <12 months) | -0.378 | 0.263 | 0.69 (0.41–1.15) | 0.151 |
| Age 24–35 months (vs <12 months) | -0.454 | 0.257 | 0.63 (0.38–1.05) | 0.077 |
| Age 36–47 months (vs <12 months) | -1.009 | 0.311 | 0.36 (0.20–0.67) | 0.001 |
| Age 48–59 months (vs <12 months) | -0.977 | 0.343 | 0.38 (0.19–0.74) | 0.004 |
| Female child (vs male) | -0.273 | 0.180 | 0.76 (0.54–1.08) | 0.129 |
| Mother’s education: primary (vs none/pre-primary) | -0.133 | 0.447 | 0.88 (0.36–2.10) | 0.765 |
| Mother’s education: secondary (vs none/pre-primary) | -0.209 | 0.436 | 0.81 (0.34–1.91) | 0.631 |
| Mother’s education: higher secondary+ (vs none/pre-primary) | -0.140 | 0.452 | 0.87 (0.36–2.11) | 0.757 |
| Poorest (vs richest) | 0.832 | 0.343 | 2.30 (1.17–4.50) | 0.015 |
| Second (vs richest) | 0.753 | 0.343 | 2.12 (1.08–4.16) | 0.028 |
| Middle (vs richest) | 0.927 | 0.331 | 2.53 (1.32–4.84) | 0.005 |
| Fourth (vs richest) | 0.548 | 0.364 | 1.73 (0.85–3.53) | 0.132 |
| Rural (vs urban) | 0.207 | 0.250 | 1.23 (0.75–2.01) | 0.408 |
| Unimproved sanitation (vs improved) | -0.267 | 0.352 | 0.77 (0.38–1.53) | 0.448 |

#### Diarrhoea models across specifications (PR, 95% CI)

| Pattern | Unadjusted | Age group and wealth only | Main model | Main + on-premises source | On-premises source only | Excluding treatment | Under-five child weight | Month covariate | Threshold ≥11 CFU/100 mL |
| --- | --- | --- | --- | --- | --- | --- | --- | --- | --- |
| ND source / Detected POU vs ND/ND | 1.09 (0.56–2.10) | 1.03 (0.53–1.97) | 1.02 (0.53–1.96) | 1.03 (0.53–1.99) | 1.35 (0.64–2.85) | 0.99 (0.52–1.88) | 1.02 (0.53–1.96) | 1.05 (0.55–2.00) | 1.06 (0.68–1.65) |
| Detected source / ND POU vs ND/ND | 1.00 (0.23–4.35) | 1.23 (0.27–5.57) | 1.20 (0.26–5.56) | 1.19 (0.26–5.54) | 1.62 (0.33–7.97) | 0.59 (0.07–4.75) | 1.21 (0.26–5.64) | 1.23 (0.26–5.78) | 3.10 (1.32–7.31) |
| Detected / Detected vs ND/ND | 1.13 (0.59–2.17) | 1.11 (0.58–2.13) | 1.10 (0.58–2.11) | 1.12 (0.58–2.18) | 1.49 (0.71–3.14) | 0.99 (0.52–1.88) | 1.10 (0.57–2.10) | 1.11 (0.58–2.13) | 1.02 (0.58–1.80) |
| POU detected vs not detected | 1.11 (0.62–1.98) | – | 1.04 (0.59–1.86) | – | – | – | 1.04 (0.58–1.85) | – | – |
| n (children) | 2,249 | 2,249 | 2,249 | 2,249 | 1,838 | 2,117 | 2,248 | 2,249 | 2,249 |

| Other diarrhoea models (n = 2,249 unless stated) | PR (95% CI) |
| --- | --- |
| Source detected vs not detected (alone) | 1.09 (0.75–1.58) |
| POU detected, adjusted for source detection | 1.01 (0.56–1.83) |
| Source detected, adjusted for POU detection | 1.09 (0.74–1.59) |
| POU detected vs not, children with a negative source (n = 1,151) | 0.97 (0.51–1.86) |

#### Diarrhoea, POU risk groups (<1, 1–10, >10 CFU/100 mL)

Children in POU-tested households, POU weight, n = 2,291 (survey design 2,442 observations); odds ratios from design-adjusted logistic regression: 1–10 CFU 1.17 (0.58–2.37); >10 CFU 1.04 (0.54–1.97). Modified Poisson PRs: 1.15 (0.60–2.22) and 1.03 (0.57–1.88).

#### Joint patterns at the >= 11 CFU/100 mL threshold (paired sample, n = 5,944; exploratory)

| Pattern | Weighted % (95% CI) |
| --- | --- |
| Both < 11 CFU/100 mL | 36.8 (35.3–38.3) |
| Source < 11 / POU >= 11 | 44.3 (42.9–45.8) |
| Source >= 11 / POU < 11 | 2.1 (1.7–2.6) |
| Both >= 11 | 16.8 (15.7–17.9) |

#### Discordance and blank-control sensitivity

Survey-weighted difference in shares (source ND / POU detected minus source detected / POU ND): 37.4 percentage points (95% CI 35.7–39.0); weighted ratio 16.0 (12.6–19.5). Excluding the 13 paired households in the seven clusters with a positive blank control: ND/Detected 39.9%, Detected/ND 2.5%, JMP-aligned 42.5%, POU-substituted 12.6%.

#### Inverse-probability-of-observation weighting sensitivity analysis

Probability of readable paired results among all POU-tested households (n = 6,106) modelled by survey logistic regression on division, residence, wealth, source type, source location and container; stabilised weights truncated at the 1st (0.98) and 99th (1.36) percentiles (untruncated range 0.98–1.62, mean 1.00) and multiplied by the water-quality weight. Assumes missingness depends only on observed characteristics.

| Estimate | Original weight, % (95% CI) | IPW-adjusted, % (95% CI) |
| --- | --- | --- |
| Source E. coli detected | 47.4 (45.9–49.0) | 47.5 (46.0–49.1) |
| POU E. coli detected | 84.8 (83.6–85.9) | 84.8 (83.7–85.9) |
| ND / ND | 12.7 (11.7–13.8) | 12.7 (11.7–13.8) |
| Source ND / POU detected | 39.9 (38.4–41.4) | 39.8 (38.3–41.3) |
| Source detected / POU ND | 2.5 (2.0–3.1) | 2.5 (2.0–3.0) |
| Detected / detected | 44.9 (43.4–46.4) | 45.0 (43.5–46.6) |
| POU detected among negative-source households | 75.8 (73.9–77.7) | 75.8 (73.9–77.6) |
| JMP-aligned safely managed estimate | 42.5 (40.8–44.1) | 42.2 (40.6–43.8) |
| POU-substituted estimate | 12.6 (11.4–13.8) | 12.5 (11.4–13.7) |
| Difference, ND/Detected minus Detected/ND (percentage points) | 37.4 (35.7–39.0) | 37.3 (35.7–38.9) |

#### Joint patterns by container at the POU test (paired sample; weighted %, 95% CI)

| Container | n | ND/ND | ND source / POU detected | Detected source / POU ND | Detected/Detected |
| --- | --- | --- | --- | --- | --- |
| Direct from source | 822 | 20.3 (17.1–23.9) | 33.3 (29.5–37.4) | 2.9 (1.7–4.7) | 43.6 (39.6–47.6) |
| Covered container | 3,911 | 11.8 (10.5–13.1) | 39.0 (37.2–40.8) | 2.3 (1.8–3.1) | 46.9 (45.0–48.8) |
| Uncovered container | 1,073 | 10.3 (8.3–12.8) | 48.3 (44.9–51.7) | 2.4 (1.5–3.9) | 39.0 (35.7–42.3) |

#### Discordance by main source type (paired sample; weighted %, 95% CI)

| Source type | n | ND source / POU detected | Detected source / POU ND | POU detected among source-negative households |
| --- | --- | --- | --- | --- |
| Piped | 1,154 | 36.4 (33.3–39.6) | 4.4 (3.2–6.1) | 66.1 (61.6–70.4), n = 617 |
| Deep tube well | 1,461 | 42.0 (39.1–45.0) | 1.0 (0.5–1.9) | 82.8 (79.3–85.8), n = 741 |
| Shallow tube well | 3,156 | 41.2 (39.1–43.2) | 2.0 (1.5–2.7) | 78.3 (75.8–80.6), n = 1652 |
| Other | 173 | 31.0 (23.2–40.1) | 5.1 (2.3–11.3) | 74.5 (58.8–85.7), n = 53 |

#### Sensitivity to the source of the glass (paired sample)

The recorded source of the glass differs from the main source code in 17.3% (weighted; 981 households). Delivered or packaged water was used by 1.1%.

| Estimate | All paired | Glass source matches main source (n = 4,963) | Excluding delivered/packaged water (n = 5,892) |
| --- | --- | --- | --- |
| Source E. coli detected, % | 47.4 (45.9–49.0) | 47.6 (45.9–49.3) | 47.5 (46.0–49.1) |
| POU E. coli detected, % | 84.8 (83.6–85.9) | 84.9 (83.7–86.1) | 84.9 (83.7–86.0) |
| ND source / POU detected, % | 39.9 (38.4–41.4) | 39.8 (38.2–41.4) | 39.8 (38.4–41.3) |
| Detected source / POU ND, % | 2.5 (2.0–3.1) | 2.5 (2.0–3.1) | 2.5 (2.0–3.0) |
| POU detected among source-negative households, % | 75.8 (73.9–77.7) | 76.0 (73.9–77.9) | 75.9 (74.0–77.7) |
| JMP-aligned estimate, % | 42.5 (40.8–44.1) | 42.1 (40.4–43.9) | 42.7 (41.1–44.4) |
| POU-substituted estimate, % | 12.6 (11.4–13.8) | 12.5 (11.3–13.7) | 12.6 (11.5–13.9) |

#### Binary container contrast (direct-from-source combined with covered containers as reference)

Mutually adjusted PR for uncovered container versus covered or direct-from-source: 1.09 (1.02–1.17); n = 2,933. Shown only to illustrate the effect of reference-group composition.

#### Spatial analysis: cluster and district Moran’s I, household mixed model, district ecological model

Exploratory spatial analysis linking water-quality clusters to MICS 2025 GPS coordinates and Geospatial Covariates (GeoCov); see Materials and methods, “Spatial data and geospatial covariates (exploratory)” and Results, “Spatial clustering and geospatial covariates”. Coordinates are displaced within the local administrative unit; covariates are cluster- or district-level averages (ecological).

**Geospatial covariate source, resolution and vintage** (from the MICS GeoCov “Data Description” sheet):

| Covariate | Source dataset | Spatial resolution | Year |
| --- | --- | --- | --- |
| Elevation | Spatial Data Access Tool (SDAT), Oak Ridge National Laboratory | ~1 km | 2000 (static terrain layer) |
| Rainfall | Climate Hazards Group InfraRed Precipitation with Station data v3 (CHIRPS) | ~5.6 km | 2020 (closest available year to the 2025 survey) |
| Population density | Gridded Population of the World v4 (GPWv4) | ~1 km | 2020 |
| Motorised travel time to healthcare | Malaria Atlas Project | ~1 km | 2019 |
| Nightlights composite | VIIRS Day/Night Band, version 2 | ~500 m | 2023 |

**Complete detection in five districts. Barguna, Jhalokati, Kushtia, Patuakhali and Sunamganj had a design-based standard error of 0 for the district estimate, because every tested household was positive. Inverse-variance weights are undefined for a standard error of 0. The main district model therefore weights each district by the number of households tested (82 to 94 per district). Two sensitivity analyses are shown in the table below: inverse-variance weights with a large finite weight for these districts, and exclusion of these districts.**

**Exact confidence intervals for the five complete-detection districts (Clopper-Pearson exact binomial 95% CI, computed on the unweighted household count for each district). The design-based estimates in the main Results use the survey weight and design-based variance. They are not identical to these exact figures, but both methods exclude the 84.8% national rate.**

| District | n households tested | 95% exact CI for true detection rate |
| --- | --- | --- |
| Barguna | 94 | 96.2–100% |
| Jhalokati | 92 | 96.1–100% |
| Kushtia | 89 | 95.9–100% |
| Patuakhali | 92 | 96.1–100% |
| Sunamganj | 84 | 95.7–100% |

The probability of observing zero negatives across 84–94 independent tests if the true district rate equalled the national POU rate (84.8%) is approximately 2×10⁻⁷ to 9×10⁻⁷ per district, so sampling chance under a uniform national rate is not a plausible explanation. This does not rule out a non-sampling (systematic) explanation specific to these districts, such as a geographically clustered field-team or equipment-batch effect, which the data available cannot distinguish from a genuine local exposure difference (Discussion, “Geographic clustering”).

**Moran’s I (global spatial autocorrelation of POU detection).**

| Level | n | Weights | Moran’s I | Expectation | z | p |
| --- | --- | --- | --- | --- | --- | --- |
| Cluster | 3,145 | 6-nearest-neighbour | 0.160 | -0.0003 | 16.5 | <0.001 |
| District | 64 (polygon-matched) | Queen contiguity (knn fallback for unlinked polygons) | 0.310 | -0.016 | 4.0 | <0.001 |

**Household-level mixed-effects logistic model, survey-weighted (POU detection ~ cluster-level GeoCov covariates + urban/rural + random intercept for PSU, household survey weight incorporated through WeMix::mix() [26]; n = 6,048 households, 3,145 PSUs). Continuous covariates standardised (z-scores).**

| Term | log-odds | SE | OR (95% CI) | p |
| --- | --- | --- | --- | --- |
| Elevation (z) | -0.472 | 0.059 | 0.62 (0.56–0.70) | <0.001 |
| 2020 annual rainfall (z) | -0.046 | 0.051 | 0.96 (0.86–1.05) | 0.360 |
| 2020 population density (z) | -0.116 | 0.071 | 0.89 (0.77–1.02) | 0.102 |
| 2019 motorised travel time to healthcare (z) | 0.503 | 0.126 | 1.65 (1.29–2.12) | <0.001 |
| Nightlights composite (z) | -0.196 | 0.082 | 0.82 (0.70–0.97) | 0.016 |
| Urban (vs rural) | -0.292 | 0.149 | 0.75 (0.56–1.00) | 0.050 |

Random-effects variance: PSU 2.895 (logistic scale); approximate ICC (PSU) = 2.895 / (2.895 + π²/3) ≈ 0.47.

**District-level ecological model (linear regression of the design-based district POU detection estimate on the same covariates; n = 67 district-level domains, comprising 64 districts and three city corporations; weighted by the number of households tested). Estimates are changes in the proportion of households with POU detection per unit of each covariate.**

| Term | Estimate | SE | t | p |
| --- | --- | --- | --- | --- |
| Elevation | -0.0012 | 0.00048 | -2.52 | 0.014 |
| 2020 annual rainfall | -1.2e-05 | 1.4e-05 | -0.87 | 0.386 |
| 2020 population density | 3.2e-06 | 4.2e-06 | 0.78 | 0.440 |
| 2019 motorised travel time to healthcare | 0.00056 | 0.00053 | 1.05 | 0.297 |
| Nightlights composite | -0.0199 | 0.0088 | -2.25 | 0.028 |

Overall model: F(5,61) = 5.67, p < 0.001, R² = 0.317, adjusted R² = 0.261. Elevation (p = 0.014) and nightlights (p = 0.028) were individually significant. The model is exploratory and ecological, so these associations do not describe individual households.

Sensitivity of the district model to weighting (overall test and individual p-values).

| Specification | n | Overall F-test p | Adjusted R² | Elevation p |
| --- | --- | --- | --- | --- |
| Weighted by households tested (main) | 67 | <0.001 | 0.261 | 0.014 |
| Inverse-variance weights, finite weight (1×10⁶) for complete-detection districts | 67 | 0.048 | 0.095 | 0.510 |
| Inverse-variance weights, five complete-detection districts excluded | 62 | 0.002 | 0.220 | 0.335 |
| Unweighted | 67 | <0.001 | 0.256 | 0.015 |

**District detection range. Design-based district POU detection ranged from complete (100%) detection in five districts (Barguna 94/94, Jhalokati 92/92, Kushtia 89/89, Patuakhali 92/92 and Sunamganj 84/84 valid household tests, all positive) to 53–65% in the lowest-detection districts. The zero standard error in these districts reflects zero observed variance among tested households.**

**Software. R 4.6.1; packages sf, spdep (Moran’s I, contiguity and k-nearest-neighbour weights) [24,25], WeMix (survey-weighted mixed model) [26], haven, survey and readxl. District boundaries: geoBoundaries BGD ADM2, CC BY 3.0 IGO [28]. Odds ratios from the mixed model overstate prevalence ratios when detection is common and are interpreted for direction only [27].**

Diarrhoea sensitivity analysis: survey month, handwashing place and treatment (n = 2,229 children). Adding these three covariates to the main model addresses season and other routes of transmission. The POU prevalence ratio was 1.04 (0.58 to 1.87), against 1.04 (0.59 to 1.86) in the main model. Joint patterns against ND/ND gave 1.04 (0.55 to 1.98) for ND source / Detected POU, 1.20 (0.26 to 5.54) for Detected source / ND POU and 1.09 (0.57 to 2.12) for Detected/Detected.

#### Minimum detectable prevalence ratios

Design effect for the weighted diarrhoea prevalence: 1.30, estimated from the survey-weighted proportion (linearised variance). Effective sample sizes after dividing by the design effect; 80% power, two-sided alpha = 0.05.

| Comparison | Reference prevalence | Minimum detectable PR |
| --- | --- | --- |
| ND/ND vs ND/Det PR | 5.5% | 2.23 |
| POU ND vs Det PR | 5.5% | 2.07 |
