## Supplementary material for "Discordance between source and point-of-use drinking-water *Escherichia coli* contamination in Bangladesh: a cross-sectional analysis and implications for safely managed water monitoring": S1_STROBE_Checklist.docx

### S1 STROBE Checklist. STROBE checklist for cross-sectional studies

Section references are to the current manuscript.

| # | Item | Where addressed |
| --- | --- | --- |
| 1a | Design in title/abstract | Title; Abstract (Materials and methods: secondary cross-sectional analysis of MICS 2025) |
| 1b | Informative abstract | Abstract |
| 2 | Background/rationale | Introduction |
| 3 | Objectives | Introduction (primary aim; secondary and exploratory aims) |
| 4 | Key elements of design | Materials and methods: Data and study design; Ethics statement |
| 5 | Setting, locations, dates | Materials and methods: Data and study design; Ethics statement; Water testing (24 February–30 June 2025; two tests dated July) |
| 6a | Eligibility, sources, selection | Materials and methods: Data and study design; Ethics statement; Results: Sample (S2 Fig) |
| 7 | Variables: outcomes, exposures, confounders | Materials and methods: Water testing; Joint status; Exposure and covariate definitions; S1 Fig |
| 8 | Data sources/measurement | Materials and methods: Water testing (membrane filtration, blanks), Exposure and covariate definitions |
| 9 | Bias | Materials and methods: Statistical analysis (minimal versus mutual adjustment; selection on negative source; sensitivity analyses with stated threats); Discussion: Source and point-of-use detection, and their discordance; Strengths and limitations |
| 10 | Study size | Results: Sample; Methods (minimum detectable PR) |
| 11 | Quantitative variables | Materials and methods: Water testing; Exposure and covariate definitions |
| 12a | Statistical methods incl. confounding | Materials and methods: Statistical analysis |
| 12b | Subgroups, interactions | Results: Factors associated with POU detection (interaction); Tables 2 and 5 |
| 12c | Missing data | Materials and methods: Statistical analysis (complete case); Results: Sample; S2 File |
| 12d | Sampling strategy/weights | Materials and methods: Statistical analysis (PSU, stratum, weights) |
| 12e | Sensitivity analyses | Materials and methods: Statistical analysis; Results; S1 File |
| 13a | Numbers at each stage | Results: Sample; S2 Fig |
| 13b | Reasons for non-participation | Results: Sample (unreadable results; missing covariates) |
| 13c | Flow diagram | S2 Fig |
| 14a | Participant characteristics | Table 1 |
| 14b | Missing data per variable | S2 File |
| 15 | Outcome data | Tables 2, 3 and 6 |
| 16a | Unadjusted and adjusted estimates | Tables 4–6 (unadjusted and adjusted PRs with 95% CI); S1 File |
| 16b | Category boundaries | Materials and methods: detection ≥1 and ≥11 CFU/100 mL; risk groups |
| 16c | Relative to absolute risk | Results: Factors associated with POU detection (adjusted prevalence differences in percentage points); S1 File |
| 17 | Other analyses | Results: Safely managed drinking water; Divisional patterns; sensitivity analyses |
| 18 | Key results | Discussion: Principal findings |
| 19 | Limitations | Discussion: Strengths and limitations |
| 20 | Interpretation | Discussion |
| 21 | Generalisability | Discussion: Strengths and limitations (time and place: February to June, one country) |
| 22 | Funding | Submission form (Financial disclosure) |
