## Supplementary material for "Discordance between source and point-of-use drinking-water *Escherichia coli* contamination in Bangladesh: a cross-sectional analysis and implications for safely managed water monitoring": S2_File_missing_data_audit.docx

### S2 File. Sample and missing-data audit

#### A. Numbers at each step

| Analysis | Eligible | Analysed | Excluded | Main reason |
| --- | --- | --- | --- | --- |
| Households sampled for water-quality testing | 6,298 | 6,106 | 192 | not occupied or test not completed (report Table SR.1.1) |
| POU results readable | 6,106 | 6,048 | 58 | unreadable or lost |
| Source samples collected | 6,106 | 6,010 | 96 | no water at source (30), source too far (2) or not recorded (64) |
| Source results readable | 6,010 | 5,999 | 11 | unreadable or lost |
| Paired, both readable (analytic sample) | 6,106 | 5,944 | 162 | no readable source or POU result |
| Negative-source households | 5,944 | 3,063 | – | selection by design |
| Table 4 complete cases | 3,063 | 2,933 | 130 | missing covariates (below) |
| Children under five in tested households | 2,442 | 2,377 in paired households | 65 | household not paired |
| Children classified as aged 0–59 months | 2,377 | 2,251 | 126 | under-five questionnaire not completed; no age recorded |
| Children with a valid diarrhoea answer | 2,251 | 2,250 | 1 | diarrhoea question not answered |
| Table 6 complete cases | 2,250 | 2,249 | 1 | missing mother’s education |

Among the 6,106 POU-tested households (POU weight), households lost from the paired sample differed from included households by division (design-based Pearson F test p < 0.001) but not by residence (p = 0.91) or wealth (p = 0.16).

#### B. Missing values by variable

**Table 4 analysis (negative-source households; base n = 3,063)**

| Variable | Missing n | Missing % |
| --- | --- | --- |
| Storage container | 70 | 2.3 |
| Treatment | 5 | 0.2 |
| Handwashing place | 39 | 1.3 |
| Collection time | 15 | 0.5 |
| Wealth | 0 | 0.0 |
| Residence | 0 | 0.0 |
| Division | 0 | 0.0 |
| Head’s education | 2 | 0.1 |
| Source type | 0 | 0.0 |
| Sanitation | 0 | 0.0 |

**Table 6 analysis (children; base n = 2,250)**

| Variable | Missing n | Missing % |
| --- | --- | --- |
| Child age | 0 | 0.0 |
| Child sex | 0 | 0.0 |
| Mother’s education | 1 | 0.0 |
| Wealth | 0 | 0.0 |
| Residence | 0 | 0.0 |
| Sanitation | 0 | 0.0 |

#### C. Testing dates (weighted % of paired households)

February 5.2; March 29.2; April 26.6; May 29.1; June 9.8; July <0.1. Source / POU detection by period (weighted %): February–March 41.3 / 80.7; April 50.1 / 85.4; May 51.2 / 86.8; June–July 50.7 / 91.8.

#### D. Blank controls

605 blank tests (one household in every fifth cluster): 598 with no E. coli, 7 with E. coli detected (1.2%; counts 1, 1, 1, 1, 2, 19, 24). Seven clusters had a positive blank; 13 paired households lie in those clusters.

#### E. Included and excluded households (POU-tested households, POU weight)

Included = readable paired results (n = 5,944); excluded = the other 162 POU-tested households. Weighted column %; design-based Pearson F test p-value for each variable. Container rows: 5,806 included and 157 excluded with an observed container.

| Variable | Category | Included, % | Excluded, % | p |
| --- | --- | --- | --- | --- |
| Residence | Urban | 30.1 | 29.6 | 0.910 |
|  | Rural | 69.9 | 70.4 |  |
| Wealth quintile | Poorest | 21.0 | 25.5 | 0.163 |
|  | Second | 19.8 | 24.7 |  |
|  | Middle | 19.5 | 14.8 |  |
|  | Fourth | 19.7 | 21.4 |  |
|  | Richest | 20.0 | 13.5 |  |
| Division | Barishal | 5.3 | 5.7 | 0.000 |
|  | Chattogram | 18.1 | 26.5 |  |
|  | Dhaka | 27.3 | 12.3 |  |
|  | Khulna | 11.1 | 28.6 |  |
|  | Mymensingh | 7.4 | 12.7 |  |
|  | Rajshahi | 13.9 | 5.7 |  |
|  | Rangpur | 11.4 | 3.3 |  |
|  | Sylhet | 5.5 | 5.2 |  |
| Main source type | Piped | 25.6 | 26.3 | 0.000 |
|  | Deep tube well | 20.4 | 14.7 |  |
|  | Shallow tube well | 51.8 | 35.9 |  |
|  | Other source | 2.2 | 23.0 |  |
| Container at POU test | Direct from source | 14.6 | 6.7 | 0.000 |
|  | Covered container | 67.5 | 84.4 |  |
|  | Uncovered container | 17.9 | 8.8 |  |
| Source location | Source off premises | 14.8 | 38.2 | 0.000 |
|  | Source on premises | 85.2 | 61.8 |  |
