## Supplementary figures and images for "Discordance between source and point-of-use drinking-water *Escherichia coli* contamination in Bangladesh: a cross-sectional analysis and implications for safely managed water monitoring"

### S1_Fig.tif

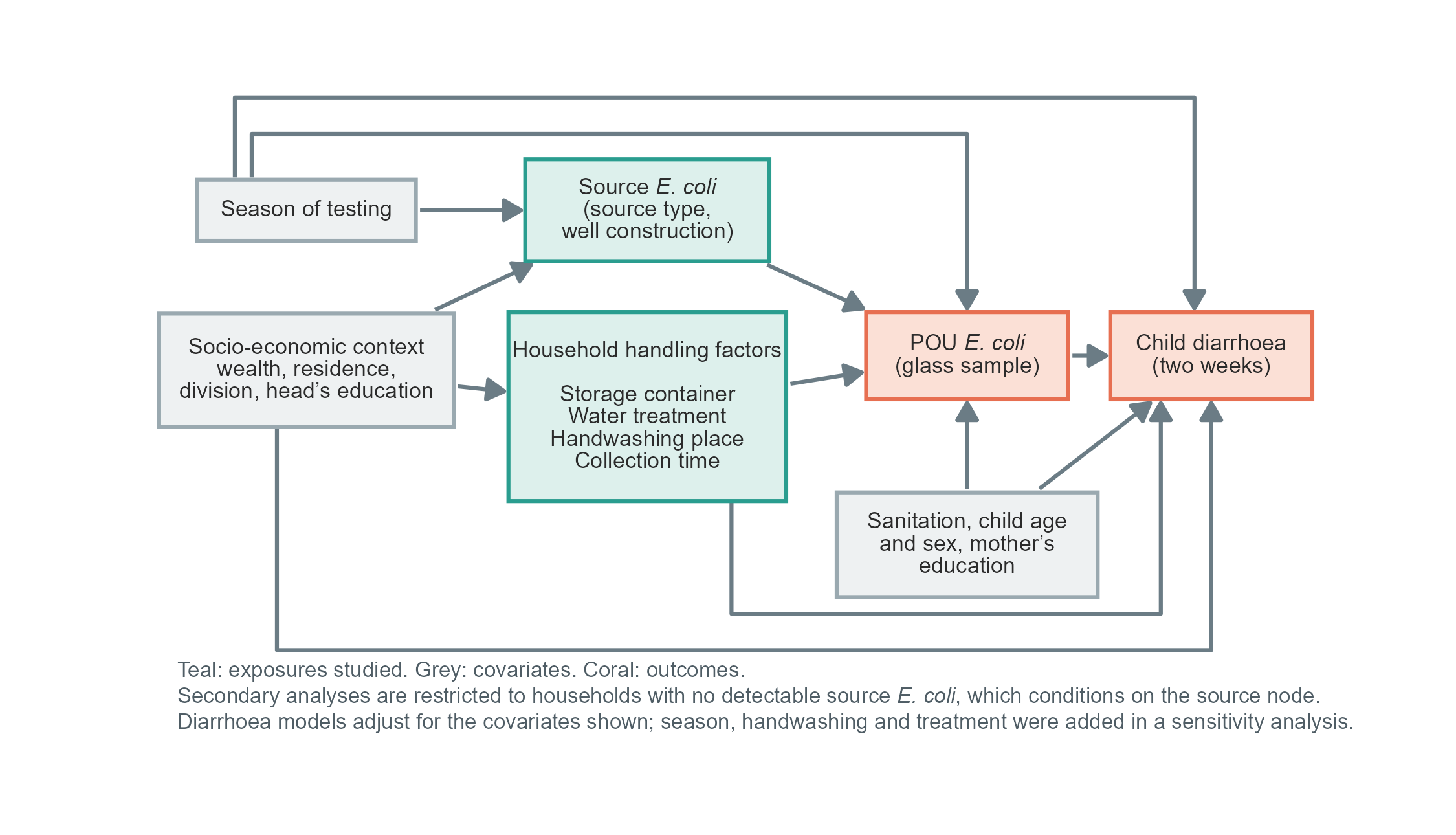

### S2_Fig.tif

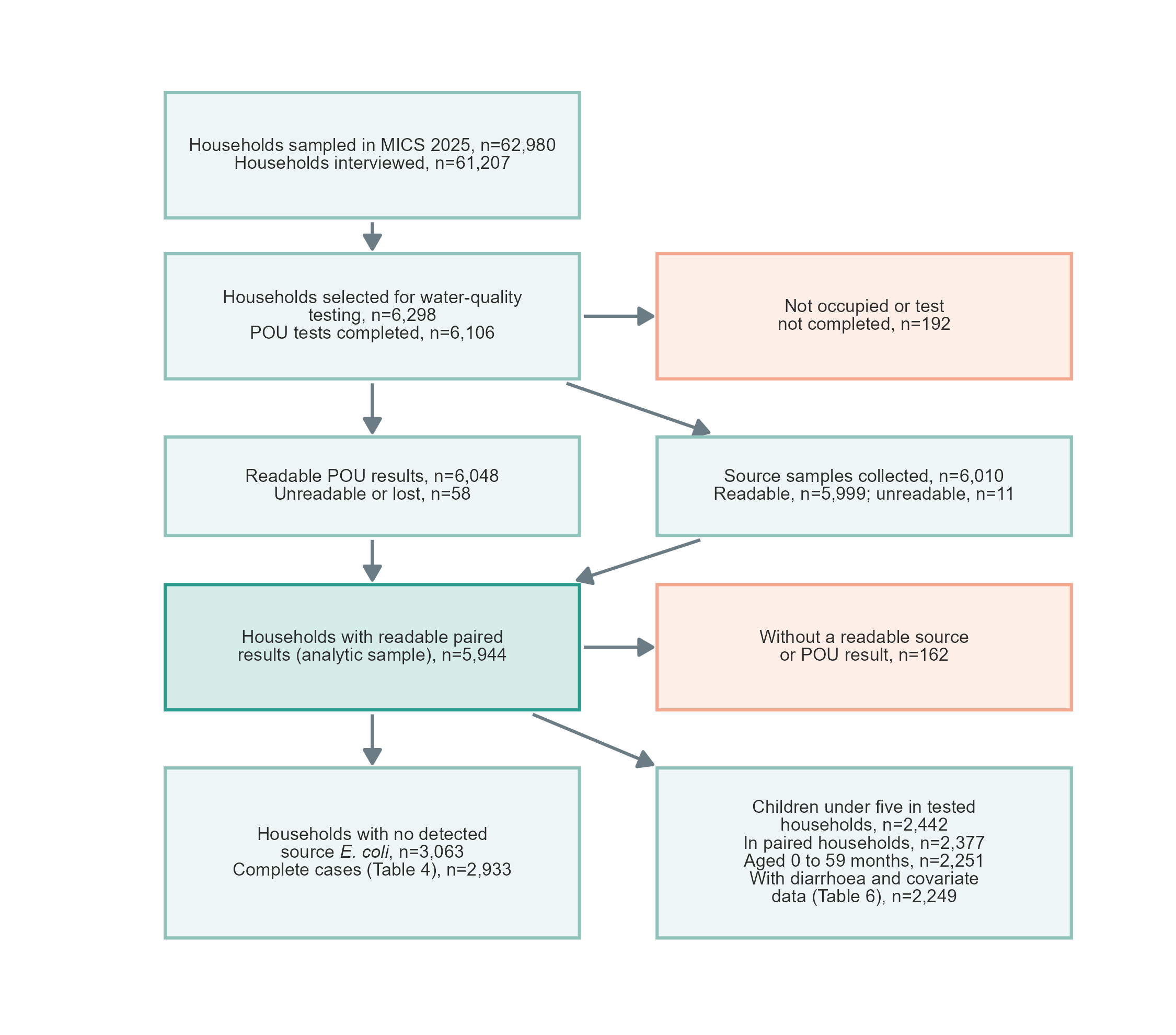
